# Dissecting Pleiotropy Between Major Depressive Disorder and Physical Disease Comorbidities

**DOI:** 10.1101/2025.10.14.25337939

**Authors:** Damian J Woodward, William R Reay, Briar Wormington, Santiago Diaz-Torres, Jue-Sheng Ong, Zachary F Gerring, Jackson G Thorp, Eske M Derks

## Abstract

Major Depressive Disorder (MDD) is characterised by substantial comorbidity with medical conditions. To achieve better outcomes for patients with MDD, an improved understanding of the mechanisms underlying pervasive comorbidities is required. To this end, we map patterns of pleiotropy by defining four clusters of physical diseases (cardiovascular, metabolic, gastrointestinal, and immune) to analyse their genetic relationship with MDD using genomic structural equation modelling. Three disease clusters exhibited independent associations with MDD and accounted for 47% of MDD *h*^2^_SNP_, with the gastrointestinal disease cluster having the strongest association (β = 0.63, SE = 0.05, *P* = 3.04 × 10^-30^). Additionally, we identified independent loci associated with the shared genetic liability between each disease cluster and MDD, revealing different pleiotropic components. Characterisation of these loci revealed previously unidentified associations with MDD and physical disease traits, along with unique biological pathways, drug groups, cell types and genes related to each disease-MDD cluster. Our findings reveal genetic connections implicating the gut-brain axis as a key mechanism underlying the comorbidity of physical diseases in MDD. This work advances our understanding of MDD by highlighting unique and shared genetic components across different disease systems.

---

Major Depressive Disorder (MDD) is characterised by considerable variation in symptoms and clinical presentations, including associations with a diverse range of medical conditions that frequently co-occur^1^. MDD ranks as the second leading cause of years lived with disability^2^, with over one-third of the total nonfatal burden in MDD patients attributed to physical disease comorbidity^3^. These comorbidities reduce adherence to medical treatment^4,5^ and increase functional disability^6^ and mortality rates^7^. At a phenotypic level, MDD is significantly associated with various conditions that can collectively be grouped into disease clusters. For example, MDD is associated with an increased risk of cardiovascular events^8,9^, metabolic dysfunction^10,11^, gastrointestinal health^12,13^, and mechanisms of the immune system^14^. The sharing of genetic risk factors may be one component underlying this relationship, with MDD demonstrating low to moderate genetic correlations across these groups of diseases^15–18^.

Here, we investigate pleiotropic components (shared genetic risk factors) between MDD and physical disease clusters and explore their downstream effects to identify genetic drivers of MDD physical disease comorbidity. We define physical diseases as non-psychiatric medical conditions and examine four physical disease clusters (cardiovascular, metabolic, gastrointestinal, and immune). These were chosen based on previous research^15,17,19,20^ which reflect common physical comorbidity groups encompassing a range of disease systems (Figure 1). We selected well-powered physical traits for each disease cluster using Linkage Disequilibrium Score Regression (LDSC), identifying 19 with a *h*^2^_SNP_ z score threshold greater than seven. Out of these traits, 16 had a significant genetic correlation with MDD (α < 2.63 × 10^-3^; 0.05/number of traits) and were retained for downstream analysis (Figure 2; Table 1). MDD had the largest correlations on average with gastrointestinal traits (mean |*r*_g_|= 0.47; range = 0.29 - 0.56), followed by cardiovascular traits (mean |*r*_g_| = 0.19; range = 0.11 - 0.29), metabolic traits (mean |*r*_g_|= 0.19; range = -0.14 - 0.23), and immune traits (mean |*r*_g_| = 0.15; range = 0.10 - 0.22) We used genomic structural equation modelling (genomic SEM)^21^ to estimate latent factors for each of the four disease clusters (representing the genetic liability that is shared across traits within each cluster) and measure their association with MDD (Supplementary Figure 1a-d, Supplementary Results Section 1.0; See Supplementary Results Section 2.0 for *h*^2^_SNP_ *Z* > 4). Each disease-latent factor had a significant association with MDD, with the gastrointestinal factor having the strongest (gastrointestinal, β = 0.69, SE = 0.03, *P* = 5.87 × 10^-158^, R^2^ = 0.48; immune, β = 0.46, SE = 0.06, *P* = 1.64 × 10^-16^, R^2^ = 0.21; cardiovascular, β = 0.31, SE = 0.02, *P* = 5.12 × 10^-46^, R^2^ = 0.09; metabolic, β = 0.23, SE = 0.02, *P* = 6.55 × 10^-44^, R^2^ = 0.05). Combining these, a four-factor model was developed to identify these factors’ independent associations with MDD (Figure 3; Supplementary Table 3; CFI = 0.918; SRMR = 0.055). When jointly modelled, we observed the gastrointestinal cluster had a strong independent association with MDD, albeit weaker than the individual model (β = 0.63, SE = 0.05, P = 3.04 × 10^-30^). The cardiovascular (β = 0.07, SE = 0.04, *P* = 0.04) and metabolic (β = -0.08, SE = 0.03, *P* = 0.02) associations with MDD were attenuated, but still demonstrated small independent associations. The immune factor had a non-significant association when controlling for the other disease clusters (β = 0.09, SE = 0.06, *P* = 0.156). Together, the four factors explained 47% of the *h*^2^ _SNP_ of MDD (R^2^ = 0.47). The combined model did not explain additional variance than the individual gastrointestinal model, indicating overlapping genetic contributions across factors. These results suggest the gastrointestinal cluster has the most significant association with MDD and largely accounts for the genetic variance of the remaining disease clusters on MDD.

**Figure 1.**
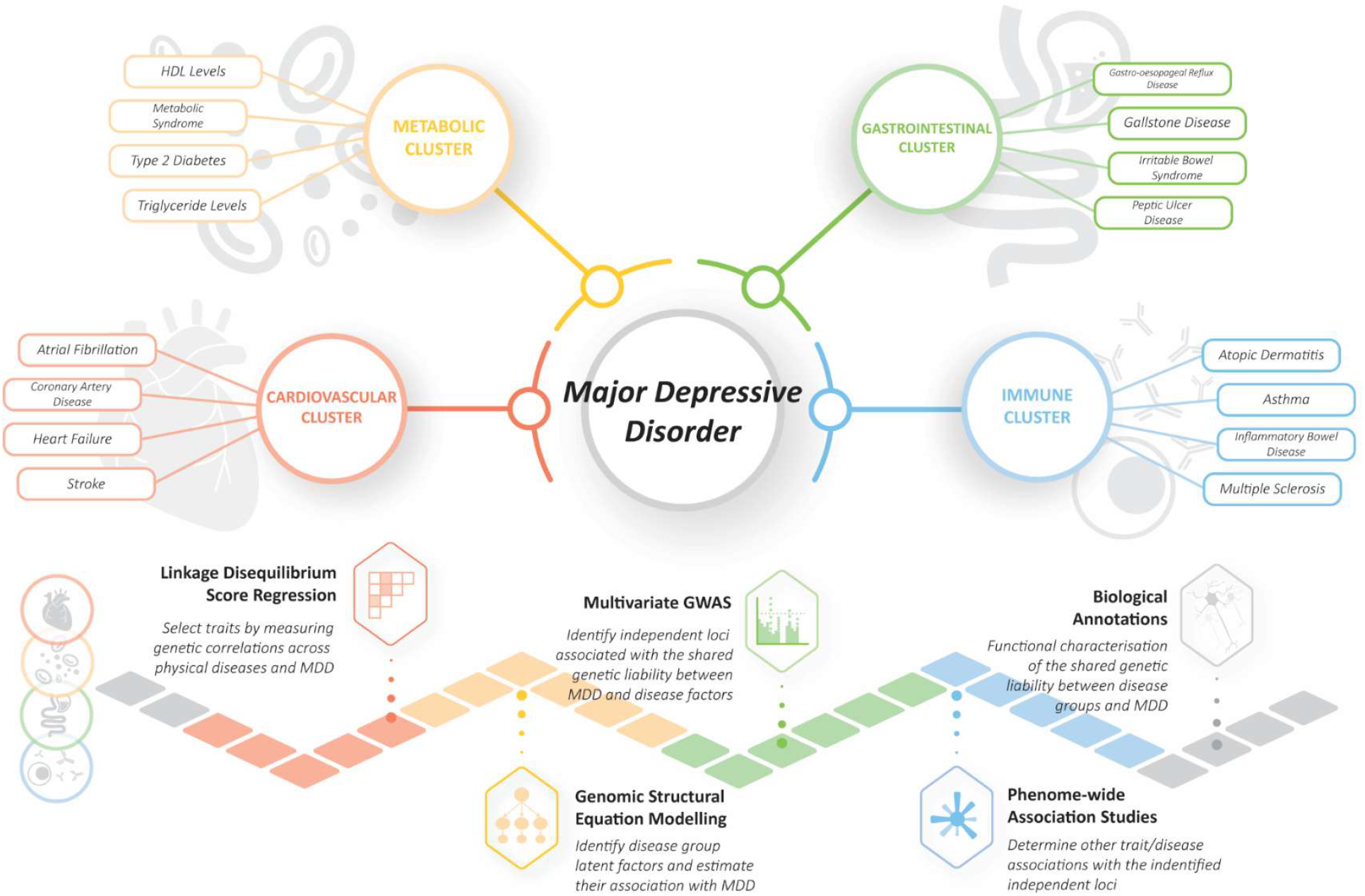
Overview figure of study design and workflow. The selected traits used for each physical disease cluster are highlighted, with each major step in the workflow outlined.

**Figure 2.**
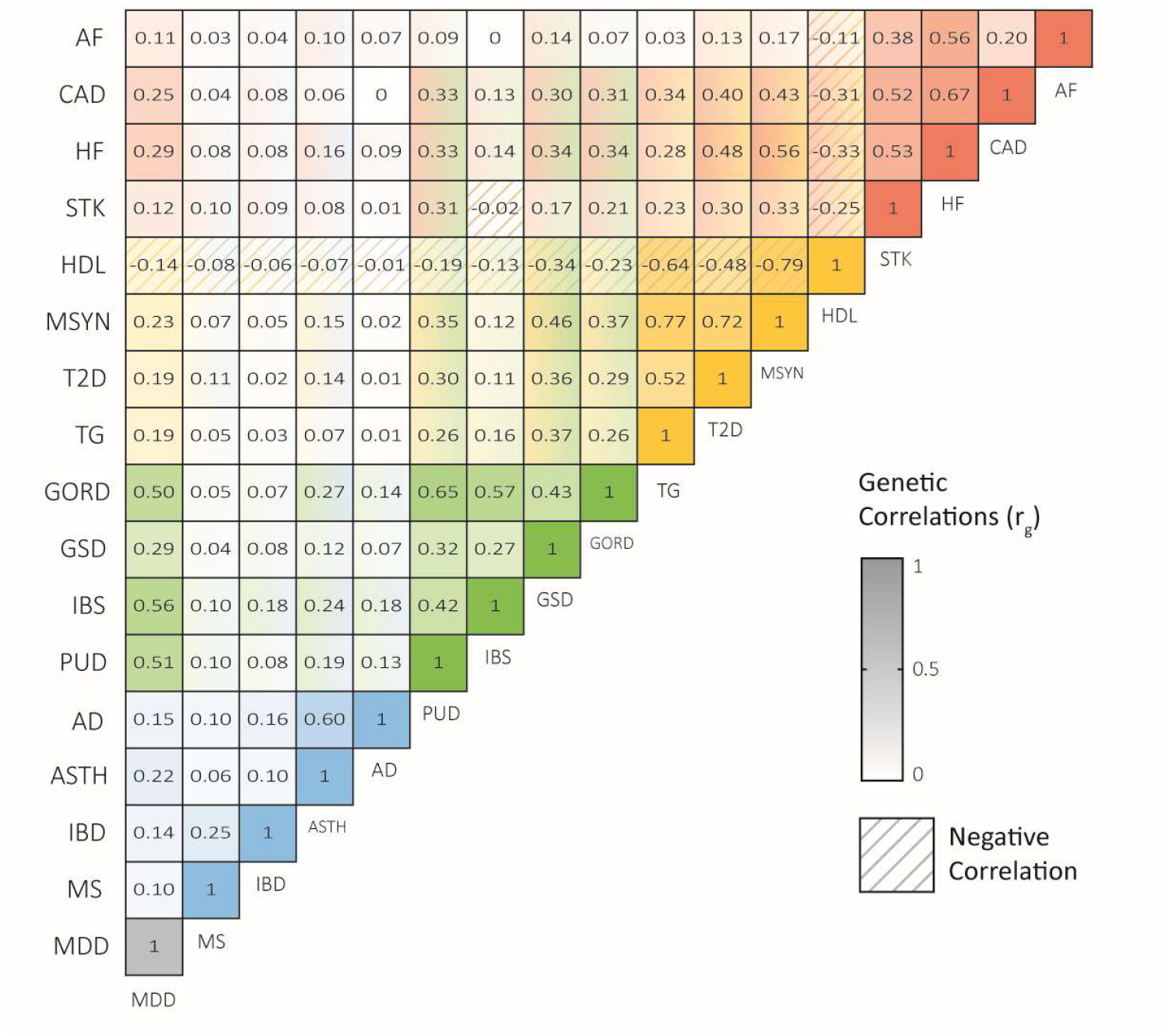
Heatmap depicting genome-wide correlations (r_g_) of disease group traits and Major Depressive Disorder (MDD) using Linkage Disequilibrium Score Regression estimated from common variants. Acronyms: Atrial Fibrillation (AF), Coronary Artery Disease (CAD), Heart Failure (HF), Stroke (STK), High-density lipoprotein cholesterol (HDL), Metabolic Syndrome (MSYN), Type 2 Diabetes (T2D), Triglycerides (TG), Gastro-oesophageal Reflux Disease (GORD), Gallstone Disease (GSD), Irritable Bowel Syndrome (IBS), Peptic Ulcer Disease (PUD), Atopic Dermatitis (AD), Asthma (ASTH), Inflammatory Bowel Disease (IBD), Multiple Sclerosis (MS).

**Table 1.** Traits identified for disease groups used for latent factor analysis and their genetic correlation (r_g_) with Major Depressive Disorder (MDD).

| Disorder/Traits | Abbreviations | Sample Size | $h^2_{\text{SNP}}$ | $h^2_{\text{SNP}} Z$ | MDD $r_g$ | $r_g P$ |
| --- | --- | --- | --- | --- | --- | --- |
| *Atrial Fibrillation* | AF | 1,030,836 | 0.0641 | 12.70 | 0.11 | $1.79 \times 10^{-07}$ |
| *Coronary Artery Disease* | CAD | 1,165,690 | 0.0541 | 17.80 | 0.25 | $1.99 \times 10^{-42}$ |
| *Heart Failure* | HF | 977,323 | 0.0288 | 12.50 | 0.29 | $4.57 \times 10^{-23}$ |
| *Stroke* | STK | 446,696 | 0.0295 | 9.04 | 0.12 | $2.51 \times 10^{-04}$ |
| *High-density Lipoprotein Cholesterol* | HDL | 1,320,016 | 0.1014 | 14.60 | -0.14 | $2.85 \times 10^{-21}$ |
| *Metabolic Syndrome* | MSYN | 291,107 | 0.1951 | 18.90 | 0.23 | $1.80 \times 10^{-27}$ |
| *Type 2 Diabetes* | T2D | 1,812,017 | 0.1286 | 22.80 | 0.19 | $2.69 \times 10^{-28}$ |
| *Triglycerides* | TG | 1,320,016 | 0.0874 | 11.00 | 0.19 | $2.32 \times 10^{-32}$ |
| *Gastro-oesophageal Reflux Disease* | GORD | 456,327 | 0.0822 | 19.60 | 0.50 | $7.26 \times 10^{-66}$ |
| *Gallstone Disease* | GSD | 550,437 | 0.0875 | 11.40 | 0.29 | $5.42 \times 10^{-32}$ |
| *Irritable Bowel Syndrome* | IBS | 486,601 | 0.0732 | 17.30 | 0.56 | $1.10 \times 10^{-83}$ |
| *Peptic Ulcer Disease* | PUD | 456,327 | 0.0725 | 8.13 | 0.51 | $9.49 \times 10^{-38}$ |
| *Atopic Dermatitis* | AD | 864,982 | 0.0551 | 7.18 | 0.15 | $1.67 \times 10^{-09}$ |
| *Asthma* | ASTH | 303,859 | 0.1434 | 13.60 | 0.22 | $1.51 \times 10^{-30}$ |
| *Inflammatory Bowel Disease* | IBD | 59,957 | 0.1517 | 11.60 | 0.14 | $1.23 \times 10^{-07}$ |
| *Multiple Sclerosis* | MS | 41,505 | 0.1149 | 11.40 | 0.10 | $2.94 \times 10^{-05}$ |

**Figure 3.**
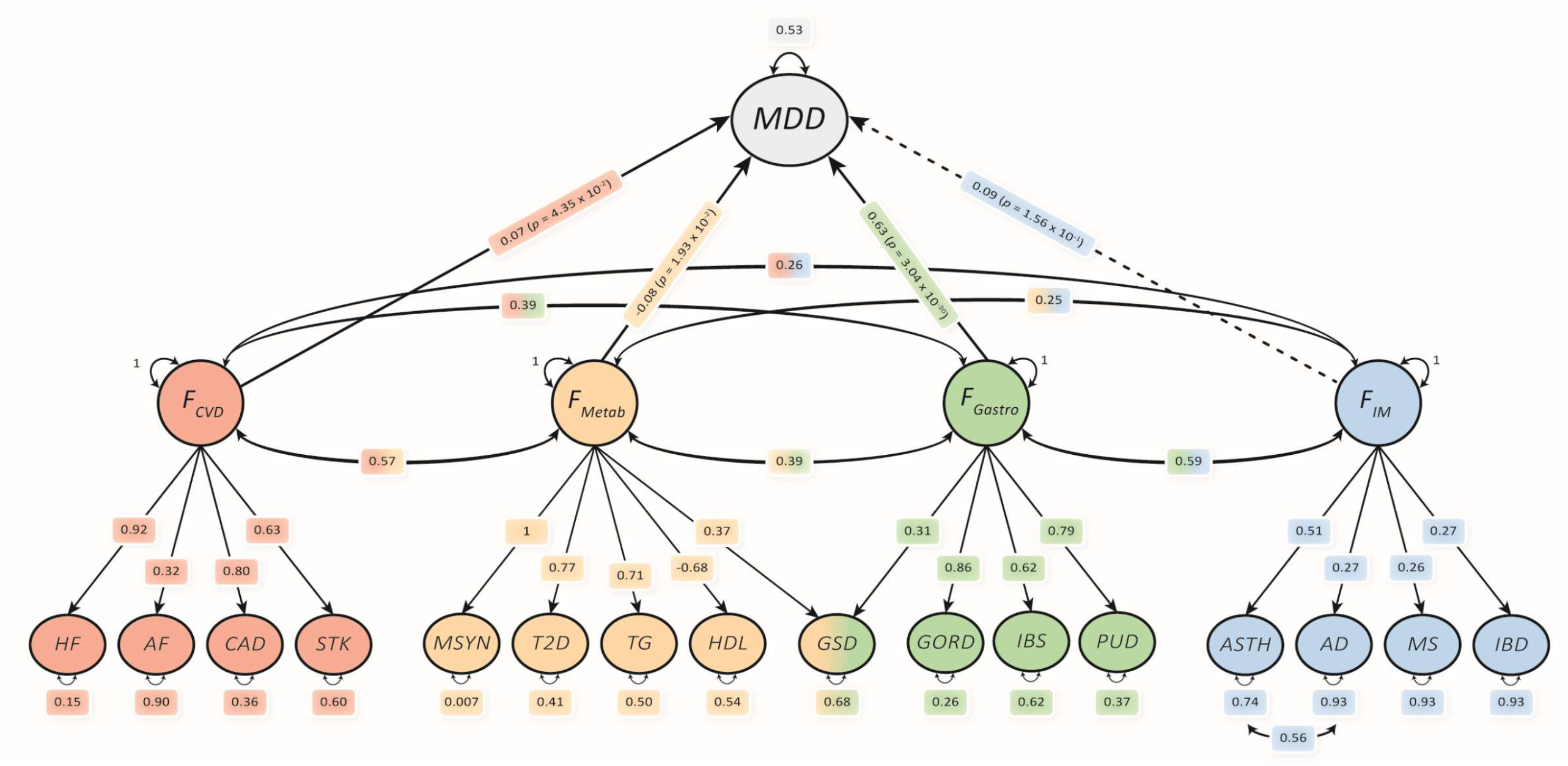
Multiple regression model of disease group latent factors (F) and their independent associations with Major Depressive Disorder (MDD) estimate using genomic structural equation modelling. A solid line demonstrates a significant independent association, while a dotted line indicates an insignificant association. White outlines represent estimated parameters. Acronyms: Cardiovascular (CVD), Metabolic (Metab), Gastrointestinal (Gastro), Immune (IM). Acronyms: Atrial Fibrillation (AF), Coronary Artery Disease (CAD), Heart Failure (HF), Stroke (STK), Metabolic Syndrome (MSYN), Type 2 Diabetes (T2D), Triglycerides (TG), High-density lipoprotein cholesterol (HDL), Gallstone Disease (GSD), Gastro-oesophageal Reflux Disease (GORD), Irritable Bowel Syndrome (IBS), Peptic Ulcer Disease (PUD), Asthma (ASTH), Atopic Dermatitis (AD), Multiple Sclerosis (MS), Inflammatory Bowel Disease (IBD).

We aimed to functionally characterise each cluster’s shared component with MDD to identify pleiotropic loci, biological mechanisms, and secondary risk pathways driving MDD physical disease comorbidity. To capture the shared genetic component between MDD and each disease cluster, we estimated models in genomic SEM in which a second-order latent factor loaded onto both MDD and the lower-order disease factor (*disease*-MDD factor) (Figure 4A; Supplementary Results, Section 3.1). We then conducted a GWAS on each disease-MDD latent factor to identify pleiotropic variants between the disease cluster and MDD. To ensure that loci associated with the disease-MDD factors are not driven primarily by a single trait, variants with highly discordant effects (Q_SNP_P < 5 × 10^-8^) were excluded (Supplementary Results, Section 3.2).

**Figure 4.**
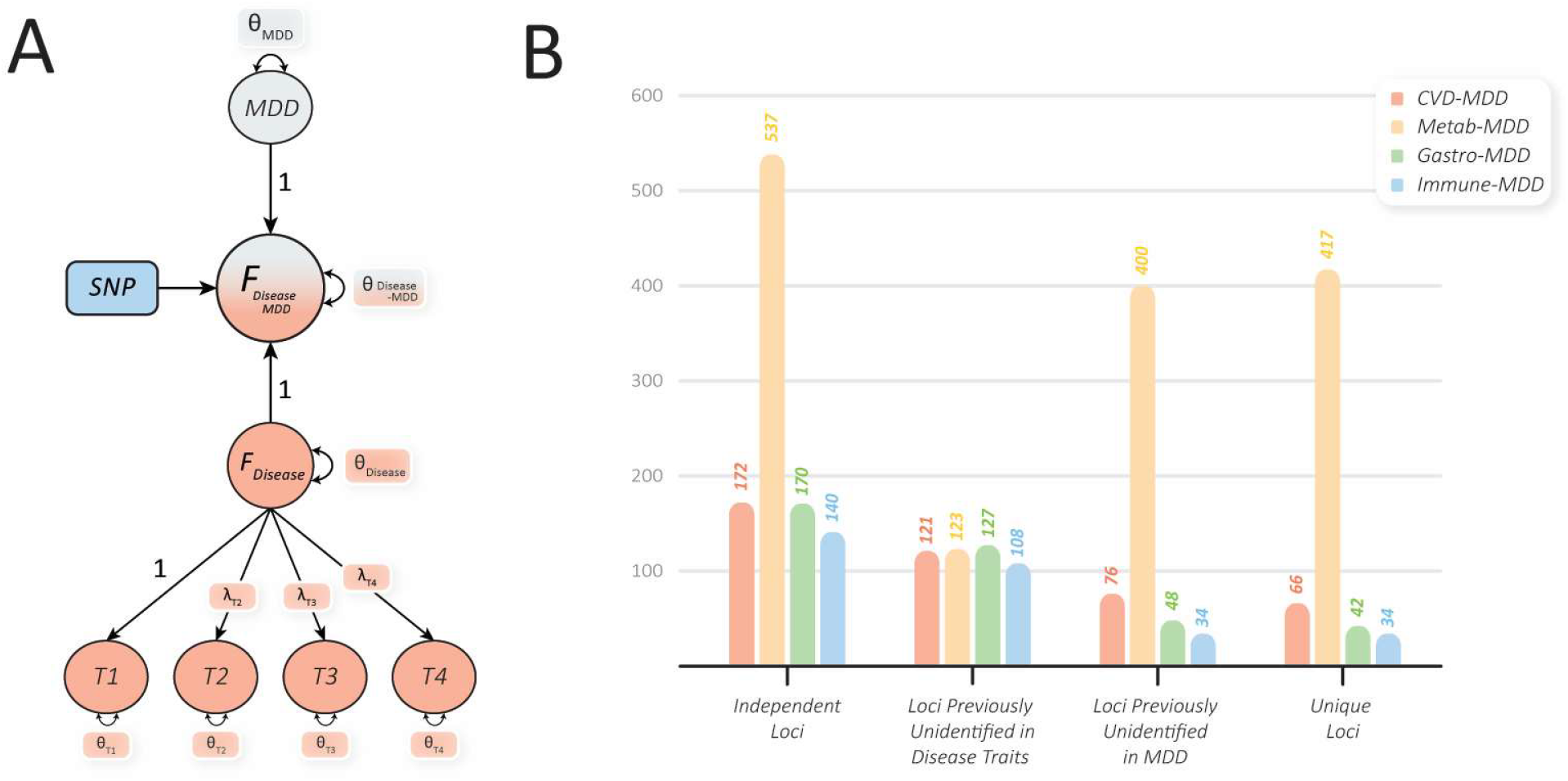
Multivariate genome-wide association study (GWAS) on second-order disease-major depressive disorder (MDD) latent factors, estimated using genomic structural equation modelling. A) Second-order factor (F_Disease-MDD_) model for multivariate GWAS capturing shared genetic liability between MDD and disease factor (F_Disease_) with unstandardised factor loadings constrained to be equal for model identification. B) Number of independent loci associated with the second-order factor disease models estimated using FUMA (r^2^ > 0.6, LD block distance > 250kb), further partitioned into the number of loci previously unidentified to disease traits, previously unidentified to MDD, and unique to the disease-MDD factor. Acronyms: Cardiovascular (CVD), Metabolic (Metab), Gastrointestinal (Gastro).

Figure 4B illustrates the independent risk loci identified from the multivariate GWAS for each disease-MDD factor (*r*^*2*^ = 0.60; maximum distance between MDD blocks ≥ 250kb; Supplementary Tables 10-13). Loci were classified into those previously unidentified for MDD and the disease cluster traits. These loci may indicate specific mechanisms more closely related to the shared genetic variance between disease clusters and MDD rather than the individual traits. Additionally, loci were compared between each disease-MDD factor to identify those unique to each factor. These loci may indicate the disease cluster-specific genetic drivers of MDD comorbidity.

First, the metabolic-MDD factor had the highest number of independent loci, with 537 associations, of which 417 were unique to this factor (not observed in the other factors). This cluster’s large number of loci highlights the strong polygenicity and statistical power of the univariate metabolic GWAS. Next, 172 independent loci were identified for the CVD-MDD factor. Of these loci, 66 were unique to the CVD-MDD factor. For the gastrointestinal-MDD factor, 170 independent loci were associated, including 42 loci unique. Lastly, the immune-MDD factor had the smallest number of independent loci, with 141. This is likely restricted by the low variance captured within the immune latent factor, as the weak genetic correlations among immune traits hindered the boost of effective sample size in this model. Of the identified risk loci in the immune-MDD factor, 34 were unique. Phenome-wide association studies (PheWASs) were also performed to determine diseases/traits associated with the independent loci using the Open Targets Genetics platform^22^. These results were largely confirmatory, with high associations observed in psychiatric traits and known risk factors for each disease cluster, supporting that the captured variance was linked to MDD and the disease cluster (Supplementary Results, Section 4.0). For example, the CVD-MDD second-order factor had high counts in anthropometric measures and psychiatric and cardiovascular traits.

Given the strong correlations between MDD and the gastrointestinal cluster, the high MDD variance explained by the gastrointestinal factor, and the high psychiatric associations observed in the PheWAS (Supplementary Results, Section 4.3), this factor was hypothesised to capture the psychophysiological mechanisms behind gut-brain-axis dysregulation. These findings are consistent with the observation that psychological factors (e.g., stress, anxiety or depression) influence gastrointestinal function and symptoms^23,24^. Importantly, some gastrointestinal traits, including GORD, can be separated into functional and non-functional subgroups. Functional GORD is a disorder subgroup associated with symptoms of GORD but with the absence of pathological acid damage along the oesophageal tract^25^. Therefore, the relationship between functional gastrointestinal disorders and the second-order factor was further investigated using GORD subgroup GWASs (Supplementary Results, Section 5.0). Overall, this follow-up analysis demonstrated that the second-order factor was primarily associated with functional GORD, indicating that the captured variance likely reflects psychophysiological mechanisms linked to the gut-brain axis rather than gastrointestinal tract pathology.

We further explored MDD and the gastrointestinal cluster to elucidate whether this strong relationship arises due to genetic risk acting through the same pathway (horizontal pleiotropy) or acting through one trait that influences the other (vertical pleiotropy) using the latent causal variable (LCV)^26^ and Causal Analysis Using Summary Effect (CAUSE)^27^ models. We found evidence for a bidirectional causal effect (vertical pleiotropy) between the MDD genetic liability and the gastrointestinal cluster, with a slightly stronger effect in the direction from MDD to the gastrointestinal cluster (See Supplementary Results for limitations, Section 5.2; Supplementary Figure 10).

Next, we performed a competitive gene-set analysis to determine biological pathways (GO gene sets) and drug Anatomical Therapeutic Chemical (ATC) codes that were enriched in the heritability of each second-order factor using GSA-MiXeR^28^ (Supplementary Results, Section 6.1). Analyses were also conducted on the univariate MDD GWAS for comparison (MDD-only). Each second-order factor demonstrated enrichment in GO gene sets related to the central nervous system that was not observed in the MDD-only analysis. Additionally, ATC drug codes associated with psychiatric treatment were found at Levels 2 and 3 of the ATC code hierarchy, supporting a psychosomatic component captured by the latent factors. The CVD and metabolic-MDD latent factors exhibited gene-set enrichment relating to cardiometabolic biological pathways. The CVD, metabolic, and gastrointestinal-MDD factors showed enrichment with ATC codes related to their respective disease clusters. The drug-gene set, propulsives, for improved gut motility, was a mutually enriched code across the MDD-only, CVD, gastrointestinal, and immune-MDD factors, supporting associations between physical disease comorbidities and MDD linked by the gut-brain axis. In addition, anxiolytics (drugs used to treat anxiety and/or depression) were observed across these second-order factors but not in MDD-only. GSA-MiXeR does not produce formal P values, so these results were compared to pre-filtered MAGMA gene-set analysis for a more conservative approach. Overall, few gene sets were observed in both analyses. MAGMA identified minimal associations, with most being larger gene sets and gene sets for the well-powered metabolic-MDD factor compared to those observed in GSA-MiXeR, highlighting the differences and limitations between the two approaches^28^.

A transcriptome-wide structural equation model was performed to determine gene expression patterns across tissues (*n* = 45) and to identify genes with expression associated with each second-order factor using genomic SEM and FUSION^29^ (Methods). Two additional analyses were performed, including an aggregated Cauchy association test^30^ (ACAT) to meta-analyse p-values and determine genes with a significant effect across tissues (ACAT hits), and colocalisation to pinpoint variants with a high likelihood (PP4 > 0.80) to be associated with the disease-MDD factor and changes in gene expression, suggesting putative causal genes (COLOC genes). The ACAT analysis identified 2,569 unique significant genes across disease-MDD factors expressed in at least one tissue, with 2,271 not identified in MDD univariate analysis (Figure 5A). We next examined the proportion of colocalised genes across tissues, normalised to z scores to compare across disease clusters. A high proportion of colocalised genes (Z > 1.96) were observed in the gastrointestinal tract, including stomach, small intestine and colon tissue in disease-MDD factors and MDD-only (Figure 5B). Additionally, the adrenal gland had a high proportion of colocalisation signal in the CVD-MDD factor, which may link to the hypothalamic-pituitary-adrenal axis, a possible shared driving mechanism of depression and CVD disease^31^.

**Figure 5.**
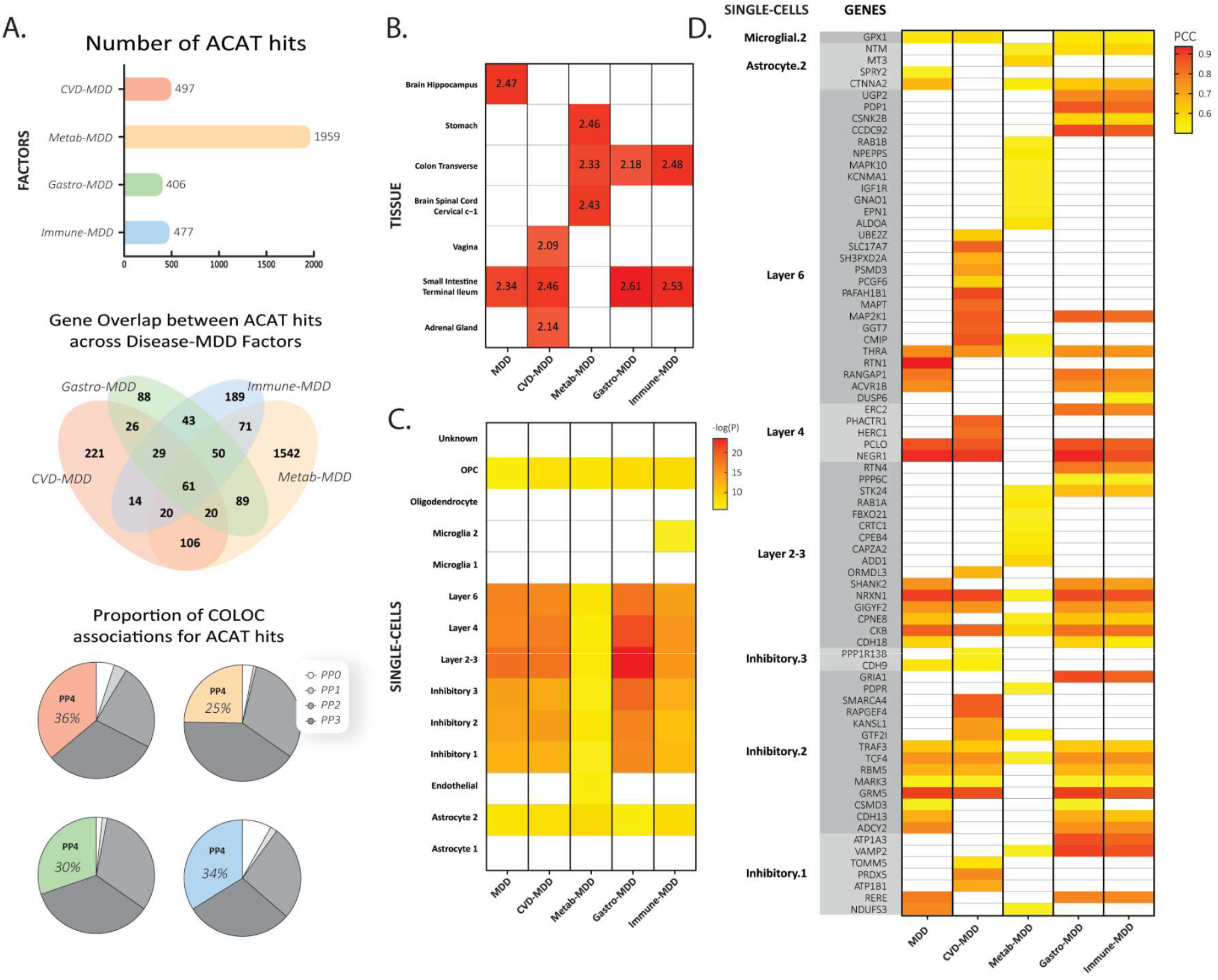
Functional annotations of second order disease-MDD factors. A) The number of aggregated Cauchy association test (ACAT) hits per tissue, the overlap across disease-MDD factors, and the proportion of coloc genes with a posterior probability (PP4) greater than 0.80. B) The high proportion of colocalised genes per tissue (coloc genes/number of genes), normalised to z scores to compare across factors (Z > 1.96). C) gsMAP results using human brain cortex tissue to identify single-cell associations across disease-MDD factors and MDD-only. D) gsMAP results using human brain cortex tissue to identify cell-type specific gene associations with each factor and MDD-only.

Lastly, spatial transcriptomics analysis was conducted using gsMAP and single-cell data from the human brain cortex. This method integrates spatial transcriptomic data with GWAS summary statistics to map cell types to complex traits and identify cell type-specific gene associations. Inhibitory cell types, astrocytes, oligodendrocyte progenitor cells and layers 2, 3, 4, and 6 of the brain cortices showed a significant signal across factors (P Cauchy < 0.05/14 - number of cell types; Figure 5C). The immune-MDD factor displayed a distinct association with the immune cell type microglia, indicating potential immune mechanisms in the brain related to the shared liability of immune diseases and MDD. Figure 5D illustrates gene associations relevant to specific cell types and latent factors. Gene associations were identified in the disease-MDD factors that were not recognised in MDD alone. These findings may point to mechanisms specific to the shared genetic liability of physical disease and MDD, with cells in layer 6 exhibiting unique gene associations across various factors.

In this study, we sought to dissect and characterise pleiotropy in MDD and comorbid physical diseases. Our analysis revealed that three disease clusters explained an independent portion of the genetic component of MDD. Notably, the gastrointestinal disease cluster had the strongest association with MDD, accounting for a substantial impact from the other disease clusters, influenced by functional gastrointestinal disorders and the gut-brain axis. Additionally, we highlighted critical pleiotropic components associated with disease-MDD latent factors, including loci, tissues, single cells, and genes, which are distinct from MDD alone. These elements may be pivotal genetic drivers of MDD-physical disease comorbidities. In line with prior research^17^, we demonstrate a shared genetic basis underlying the gut-brain axis, MDD and gastrointestinal diseases. Our results expand on these findings, showing that this is likely due to psychopathological pathways related to functional gastrointestinal diseases that impact other disease systems. The gut-brain axis has exhibited connections with CVD^32^, diabetes^33^, metabolic syndrome^34^ and immune diseases^35^. We observe that this common mechanism may influence the genetic relationship of these diseases with MDD.

Current treatments for MDD are not effective for all patients, with physical comorbidities influencing treatment outcomes^36^. However, addressing pleiotropic components may have a broader impact on comorbidities^37^. The management of MDD comorbidities has demonstrated synergistic benefits; for example, antidepressants have positively influenced physical disease outcomes^20^, such as glycaemic control in diabetics^38^. Conversely, certain physical disease treatments enhance the effects of antidepressants (e.g., anti-inflammatory medications^39^ and statins^40^). Concerning functional gastrointestinal disorders, both psychotropic and psychological interventions have proven effective in alleviating gastrointestinal symptoms^41,42^. Our findings accentuate the crucial psychopathological connections related to the gut-brain axis, presenting it as a key mechanism in physical disease comorbidities and a likely pathway affecting treatment outcomes. Considering the bidirectional nature of these conditions, treatment may require a multidisciplinary approach that encompasses addressing these pathways alongside managing physical symptoms. Future research should investigate disease causality, the impact of the gut microbiome, and the role of environmental factors such as diet to improve treatment outcomes.

The findings of this study need to be interpreted in light of the following limitations. First, our study was limited to individuals of European ancestry due to data availability. Furthermore, the current GWAS sample sizes did not allow the inclusion of other physical disease associations (e.g., immune disorders: psoriasis vulgaris, systemic lupus erythematosus^43^). Power differentials between MDD and the disease factors resulted in stronger influences in the second-order factors despite equivalent modelled variance; for instance, the metabolic-MDD factor exhibited a stronger metabolic association, leading to downstream results in the nervous system and psychiatric signal to be relatively diluted.

In conclusion, our findings highlight the shared genetic architecture between MDD and physical disease clusters. These results emphasise the complex interactions among physical disease systems and their interplay with MDD, suggesting genetic factors that may influence the management of comorbid physical diseases.

## Methods

### Data Selection and Processing

The MDD summary statistics from Als et al. (2023) (without 23andMe) were used for this analysis^44^. To identify traits to include within each disease group (cardiovascular, metabolic, gastrointestinal, and immune), we searched the published literature for disorders/traits commonly comorbid with MDD and related to each group. GWAS summary statistics of determined traits were downloaded mostly from data repositories (GWAS catalogue, National Bioscience Database Centre Human Database) (see Supplementary Table 1 for full details). Only data from the largest European ancestry studies were considered due to the limited availability of well-powered GWAS of other ancestries. Summary statistics were processed using the *munge* function within the Genomic SEM R package v0.0.5^21^ with default parameters and the 1000 Genomes European linkage disequilibrium (LD) reference panel. SNPs were filtered using an INFO filter of 0.90 and a minor allele frequency filter of 0.01 if this information was available.

### LD Score Regression

To determine which traits have significant genetic overlap with MDD, Linkage Disequilibrium Score Regression (LDSC) was performed using the *ldsc* function in Genomic SEM R package with default settings. LDSC estimates bivariate genome-wide correlations and accounts for sample overlap. Due to its complex LD structure, the MHC region was excluded from the analyses. Each trait’s SNP-based heritability was estimated within LDSC, with traits retained using two SNP-based heritability z score thresholds, *Z* > 4 and *Z* > 7. To identify MDD-related latent factors, traits that passed the threshold and were significantly correlated with MDD following correction for multiple testing (genetic covariance *P* < 0.05 /number of traits above threshold) were used for latent factor analysis.

A representative set of traits for each disease group was retained (Table 1). All groups contained four traits. The cardiovascular group includes atrial fibrillation, coronary artery disease, heart failure, and stroke. The metabolic group comprises high-density lipoprotein cholesterol, metabolic syndrome, type 2 diabetes, and triglycerides. The gastrointestinal group includes gastroesophageal reflux disease, gallstone disease, irritable bowel syndrome, and peptic ulcer disease. The immune group comprises atopic dermatitis, asthma, inflammatory bowel disease, and multiple sclerosis.

### Latent Factor Models

In all described models, parameters were estimated using diagonally weighted least squares and two indices were used to evaluate model fit. These indices included comparative fit index (CFI) and standardised root mean square residual (SRMR), using thresholds of CFI > 0.90 and SRMR < 0.10 for acceptable fit. Additionally, in instances of a Heywood case, constraints were applied to amend the discrepancy, such as fixing a negative variance to greater than zero.

#### Common Factor Models

We fitted factor models for each disease group separately to assess whether a factor could capture the shared variance across the identified traits and estimate the overall relationship between MDD and the disease clusters. The *usermodel* function in genomic SEM^21^ was used to estimate the models, using unit variance identification. For each disease group, the individual traits loaded onto a single common latent factor. MDD was then regressed onto the latent factor to estimate the disease factors’ genetic association with MDD.

#### Four-factor Multiple Regression Model

The previously identified models were combined into a four-factor model. This model was fitted in genomic SEM to determine the independent associations of each disease latent factor on the heritable component of MDD. This model also enabled the estimation of the residual variance of MDD unexplained by the four physical disease groups. MDD was simultaneously regressed onto each disease group’s latent factor, with all factors correlating with each other. Unit factor identification was used for model identification.

#### Multiple Regression of Individual Traits

A multiple regression model was specified in genomic SEM by regressing MDD onto the traits that made up the disease factors. This approach was used to identify specific traits driving the relationship between the disease factor and MDD, highlight unique associations with MDD not captured by the factor, and compare the MDD variance explained by the individual traits versus the latent factor. The traits encompassing a latent factor with a significant independent association were included in a multiple regression model.

### Multivariate GWAS

#### Second-order latent factor models

A multivariate GWAS was performed within genomic SEM for each disease group and MDD. Higher-order factor models were developed using a second-order latent factor loaded onto the disease group latent factor and MDD, with equality constraints fixed on these loadings to identify the model. This second-order factor captures the shared variance between the disease factor and MDD, enabling further characterisation of their genetic overlap and identifying variants broadly affecting a disease group and MDD. This model hypothesises that a common genetic factor mediates the relationship between MDD and the disease group and that this relationship is mediated equally. The *userGWAS* function with a diagonally weighted least squares estimator was used to conduct the multivariate GWAS. The model was identified using unit loading identification by fixing the factor loading of the trait with the largest loading from the common factor model to 1.

#### Independent Loci and Q_SNP_ Heterogeneity

Two models were produced to filter for SNP heterogeneity effects measured through Q_SNP_. Q_SNP_ is a χ^2^-distributed test statistic, with higher values indicating that a factor does not entirely explain a SNPs effect. In the first model, the SNP is loaded onto the second-order latent factor, i.e. the SNP acting through a common pathway. In the second model, the SNP is loaded onto MDD and the disease factor, representing the SNP acting through an independent pathway. The χ^2^ of the common and independent pathways were then compared to derive the Q_SNP_ heterogeneity metric. SNPs were removed if their effect was consistent with the independent pathway or if they were in LD with a significant Q_SNP_. PLINK v1.90 *clump* function was used to identify SNPs in LD with Q_SNP_ (*P*1 = 5 × 10^-8^; *P*2 = 5 × 10^-4^; *R*^2^ = 0.10; kb = 1000). This approach ensures that any identified loci are significant for the shared factor rather than for the disease group factor and MDD independently.

The summary statistics of each multivariate GWAS were uploaded to FUMA v1.5.2^45^. Independent significant loci were defined using the PLINK clumping procedure in FUMA with default settings (*r*^2^ > 0.6, LD block distance > 250kb) and the 1000 Genomes Phase 3 European reference panel. The *Open Targets Platform*^46^ was used to identify previously unidentified loci using the *Google Big Query* platform and *open targets genetics* datasets ‘variants’ and ‘variants-disease’. A locus was classified as previously unidentified if there were no disease-associated lead or tag variants within the loci for European ancestry in the Open Targets Platform and not observed in the original GWASs. This was tested for both MDD (loci previously unidentified to MDD) and the traits encompassing disease groups (loci previously unidentified to diseases). Loci unique to each disease-MDD factor were determined using the *bedtools*^47^ intersect function. An independent locus with no overlap with any locus in the other second-order factors was categorised as unique.

### Phenome-wide Association Studies

We conducted phenome-wide association studies (PheWASs) to characterise the disease-MDD loci. PheWASs investigate the associations between a wide range of phenotypes and a variant or a set of variants. Using the previously described *Open Targets Platform*^46^ datasets, known trait associations were mapped to the identified loci. Traits were categorised into their respective groups, and the number of associations counted. Unique genome-wide significant variant-trait associations of European ancestry were considered. This was performed for each disease-MDD factor, and associations were partitioned into loci previously unidentified to disease traits, previously unidentified to MDD, and loci unique to the second-order factor.

### Gastrointestinal Factor Follow-up Analysis

#### GORD Subtypes

To investigate the association between the gastrointestinal cluster and the functional subtype, we obtained GORD variants listed in Ong et al. (2022) and the gastrointestinal-MDD latent factor summary statistics^25^. Ong et al. (2022) derived GORD subtypes from variants that were depression-driven (functional-GORD associated) and BMI-driven (non-functional GORD associated). Using a Wilcoxon test, we compared the absolute effect sizes of these variants from the gastrointestinal-MDD GWAS to explore their relationship with these gastrointestinal subtypes. LDSC in the GenomicSEM R package was also implemented to estimate the genetic correlations with MDD and GORD subtypes. LDSC was performed with MDD and the other gastrointestinal traits using default settings.

#### Latent Causal Variable

Latent Causal Variable (LCV) model^26^ was used to estimate evidence of a causal relationship in either direction between the gastrointestinal common factor and MDD. This analysis models the bivariate distribution of genome-wide effects of both GWAS (MDD and gastrointestinal common factor). Bias arising due to sample overlap is mitigated in the LCV framework using the LDSC intercept. In our LCV model, we extracted ∼ 1 million common HapMap3 non-MHC variants from munged summary statistics for both traits, setting MDD as trait one and the gastrointestinal common factor as trait two. We used the recommended threshold significantly non-zero |GCP| > 0.6, to define partial genetic causality in either direction^26^.

#### CAUSE

Causal Analysis Using Summary Effect (CAUSE) model^27^ was conducted to estimate model parameters of correlated horizontal pleiotropy and direct or vertical pleiotropic effects between the gastrointestinal common factor and MDD. The CAUSE model was implemented using the CAUSE R package v1.2.0, with sample overlap directly accounted for empirically during estimation of the correlation between variants. Models were built in both directions (MDD → gastrointestinal common factor; gastrointestinal common factor → MDD). Approximately 1 million random variants from the GWAS were used to estimate nuance parameters in the model, followed by specifically using a set of clumped variants that are at least nominally associated with the exposure to perform model comparison (*P* < 0.001, *r*^2^ < 0.01, 1000 genomes phase 3 European reference panel), in line with recommendations for the CAUSE method. Three beta prior distributions were tested with asymmetric hyperparameters (necessary for the model to be identifiable) on the *q* parameter, with *q* the proportion of variants that act through correlated pleiotropy – q ∼ Beta(1,2), q ∼ Beta(1,10), and q ∼ Beta(1,100)^27^.

### Biological Annotations

#### Gene-set Enrichment

GSA-MiXeR^28^ v2.1.1, an analytical tool for exploratory gene set enrichment, was conducted for each second-order factor and MDD-only. This analysis model’s gene-set heritability enrichment enables the quantification of partitioned heritability and fold enrichment for complex traits. Gene Ontology (GO) gene sets and European 1000 genomes Phase 3 reference panel provided by GSA-MiXeR were downloaded from https://github.com/precimed/gsa-mixer. Anatomical Therapeutic Code (ATC) drug-gene sets were obtained from DrugBank^48^. Gene sets with AIC > 0 and enrichment scores > 1 were explored with analysis performed using default parameters recommended by the software. Additionally, MAGMA gene-set analysis^49^ was performed alongside GSA-MiXeR as a conservative comparison. While GSA-MiXeR alone has shown to be stable in gene-set rankings, this method does not produce formal p-values; therefore, it relies on MAGMA as an additional step to prefilter the gene-sets using Bonferroni multiple testing correction (0.05/number of gene-sets). We report both exploratory (GSA-MiXeR without prefiltering) and conservative (GSA-MiXeR with MAGMA) approaches.

#### Gene Expression of Second-order Factors

Transcriptome-wide Structural Equation Modelling (T-SEM) was performed to determine tissue-specific imputed gene expression patterns of second-order latent factor models. First, univariate transcriptome-wide association studies were implemented on each trait using the FUSION^29^ software to impute genetically regulated gene expression using default settings. Forty-five European-specific tissue weights derived from the Genotype-Tissue Expression Program (GTEx v8)^50^ were downloaded from the FUSION website, which excluded non-natural tissues (cell cultures), tissues < 100 samples, and an expression outlier (testis tissue^51^). Second, the T-SEM function in the GenomicSEM R package v0.0.5 was implemented for this analysis. The previously determined LDSC output and second-order factor models from multivariate GWAS were used. Like Q_SNP_, a Q_GENE_ is a χ^2^-distributed test statistic calculated to filter out genes not operating through the common pathway model, removing genes with a P value < 0.05/number of genes per tissue. Aggregated Cauchy association test R package v0.91^30^ was conducted to meta-analyse p-values and identify tissues with a significant effect across tissues (ACAT hits; P > 0.05). Colocalisation was performed on each second-order factor GWAS, using the FUSION software and the same tissue weights as the previous analysis. The coloc results of this analysis were integrated with the T-SEM results to determine genes with a high probability (PP4 > 0.80) to be associated with changes in gene expression and the disease-MDD factor, indicating that a gene is putatively causal. Z scores for colocalisation gene proportions (i.e., COLOC genes/total genes in tissue) were calculated to normalise and compare colocalisation signals across factors.

#### gsMAP

Genetically informed spatial mapping of cells for complex traits (gsMAP)^52^ was conducted integrating spatial transcriptomic data with GWAS summary statistics to identify cell types and single cell-specific genes associated with each second-order factor and MDD. We used spatial transcriptomic data from the human brain frontal cortex obtained from the CosMx Spatial Molecular Imager (SMI) profiling over 6,000 genes at single-cell and subcellular resolution from formalin-fixed paraffin-embedded (FFPE) human brain tissue^53,54^. Cell types were determined using the Cauchy combination results, highlighting specific cell types for each factor and MDD (P < 0.05/14). We prioritise cell type-specific genes based on gsMAP Pearson correlation coefficient (PCC > 0.50) and MAGMA p-values using gene mapping in FUMA^45^ (window = 10kb; P < 0.05/19154). PCC summarises the relationship between spatial gene expression in cell types and their genetic enrichment. MAGMA is an additional layer of evidence supporting the gene as highly relevant to the disease.

## Supporting information

supplementary_results

## Data Availability

The project used publicly available data. The MDD GWAS summary statistics (excluding 23andMe) were downloaded from https://ipsych.dk/en/research/downloads/. See Supplementary Table 1 for download links to publicly available physical disease GWAS summary statistics. MS was obtained from https://imsgc.net/ upon request. The Open Targets Platform datasets were accessed via Google BigQuery (https://platform-docs.opentargets.org/data-access/google-bigquery). GO gene sets were downloaded from https://github.com/precimed/gsa-mixer, and ATC codes gene sets from https://go.drugbank.com/. EUR GTEx v8 tissue weights are available from the FUSION website (http://gusevlab.org/projects/fusion/). The single-cell spatial dataset of the formalin-fixed paraffin-embedded Human Frontal Cortex was obtained from nanoString (https://nanostring.com/products/cosmx-spatial-molecular-imager/ffpe-dataset/human-frontal-cortex-ffpe-dataset/).

https://imsgc.net/

https://platform-docs.opentargets.org/data-access/google-bigquery

https://github.com/precimed/gsa-mixer

https://go.drugbank.com/

http://gusevlab.org/projects/fusion

https://nanostring.com/products/cosmx-spatial-molecular-imager/ffpe-dataset/human-frontal-cortex-ffpe-dataset/

## Acknowledgements

EMD is supported by an Australian National Health and Medical Research Council (NHMRC) L1 Investigator Grant (2026364). JGT is supported by an Australian NHMRC EL1 Investigator Grant (2027002). ZFG is supported by an Australian NHMRC EL1 Investigator Grant (2034743). WRR is supported by an Australian NHMRC EL1 Investigator Grant fellowship (2025671). JSO acknowledges the Australian NHMRC Investigator Grant (GNT2018420). We acknowledge the International Multiple Sclerosis Genetics Consortium for supplying data for this project.

