## supplementary_results for "Dissecting Pleiotropy Between Major Depressive Disorder and Physical Disease Comorbidities"

### 1.0 Disease-MDD Latent Factor Analysis

Each disease factor model had their representative traits loaded onto a latent factor with MDD regressed onto that factor (Supplementary Figure 1a-d; see Supplementary Table 2 for complete model outputs). Each model used unit variance identification for model identification.

**1.1 Cardiovascular Disease Latent Factor.** The cardiovascular disease factor model contained heart failure (HF), coronary artery disease (CAD), atrial fibrillation (AF), and stroke (STK). The model fit parameters were CFI = 0.924 and SRMR = 0.067, indicating an acceptable fit. Factor loadings were strong-moderate (Mean |λ| = 0.69, Range |λ| 0.98 – 0.42). HF had the strongest standardised factor loading of 0.98, followed by CAD (λ = 0.69), STK (λ = 0.66), and lastly AF (λ = 0.42). The cardiovascular common factor was significantly associated with MDD (β = 0.31, $P$ = 5.12 x $10^{-46}$). As the residual variance of MDD was estimated to be 0.91, this disease cluster explained 9% of the genetic component of MDD (calculated as $R^2$ = 1 – 0.91).

**1.2 Metabolic Disease Latent Factor.** The metabolic factor included four traits: type 2 diabetes (T2D), metabolic syndrome (MSYN), triglycerides (TG), and high-density lipoprotein (HDL). Two Heywood cases were observed due to the MSYN trait, a standardised factor loading greater than one and negative residual variance. Therefore, two restraints were applied to fix the MSYN factor loading to be less than one and the residual variance greater than 0.001. This resulting model fit well (CFI = 0.984; SRMR = 0.030). The standardised factor loadings of this disease cluster were the strongest out of the four groups (Mean |λ| = 0.80, Range |λ| 1– 0.69). MSYN had the largest factor loading of 1. HDL had the only negative factor loading across clusters, with -0.73. TG and T2D also had high loading with 0.79 and 0.69, respectively. The metabolic disease factor was significantly associated with MDD (β = 0.23, $P$ = 6.55 x $10^{-44}$) and explained 5% of MDD genetic variance ($R^2$ = 0.05).

**1.3 Gastrointestinal Disease Latent Factor.** Gastro-oesophageal reflux disease (GORD), irritable bowel syndrome (IBS), gallstone disease (GSD), and peptic ulcer disease (PUD) were used for this disease latent factor. This model demonstrated a good fit with a CFI of 0.978 and SRMR of 0.049. The model's factor loadings were strong (Mean |λ| = 0.67, Range |λ| 0.78 – 0.44). GORD had the highest standardised factor loading with 0.78, followed by IBS (λ = 0.74), PUD (λ = 0.73), and lastly GSD (λ = 0.44). The gastrointestinal disease factor had the strongest association with MDD (β = 0.69, $P$ = 5.87 x $10^{-158}$). The factor explained the most significant proportion of genetic variance of MDD at 48%.

**1.4 Immune Disease Latent Factor.** The immune disease factor model contained four traits. These included asthma (ASTH), atopic dermatitis (AD), multiple sclerosis (MS), and irritable bowel syndrome (IBD). Due to ASTH and AD's strong correlation with each other, high residual correlation, and previous reporting as an immune-cluster subtype[1], these two traits had a fixed correlation within the model ($r_g$ = 0.53), improving model fit from CFI = 0.860 and SRMR = 0.071 to CFI = 0.955 and SRMR = 0.047. The factor loadings for these traits were the smallest out of the MDD-disease latent factor models (Mean |λ| = 0.35, Range |λ| 0.27 – 0.44). ASTH had the largest factor loading with 0.44, followed by AD (λ = 0.35), IBD (λ = 0.33), and lastly MS (λ = 0.27). The immune disease factor had a significant association (β = 0.46, $P = 1.64 \times 10^{-16}$) and explained 21% of the genetic component of MDD ($R^2$ = 0.21).

**1.5 Four Factor Model.** A four-factor model was developed, combining the four identified MDD-disease factor models (Figure 3; Supplementary Table 3). MDD was regressed onto each factor to determine the independent associations of each factor. All factors correlated with each other to account for genetic overlap between factors. The resulting model fit was acceptable using the same constraints, traits, and parameters as the individual models (CFI = 0.903; SRMR = 0.065). GSD had a high level of residual covariance across the metabolic traits (residual correlation > 0.10); therefore, GSD was loaded onto both metabolic (λ = 0.37) and gastrointestinal disease factors (λ = 0.31), improving model fit (CFI = 0.918; SRMR = 0.055).

The proportion of MDD $h^2_{SNP}$ explained by the four factors was 47% ($R^2$ = 0.47). The cardiovascular disease factor had a significant independent association with MDD (β = 0.07, $P$ = 0.044), with the strength reduced from 0.31 when accounting for the other factors. The cardiovascular correlation with the metabolic disease factor was the highest at 0.57, followed by the gastrointestinal factor ($r_g$ = 0.39) and the immune factor ($r_g$ = 0.26). The metabolic factor was significantly associated with MDD (β = -0.076, $P$ = 0.019). This was the only negative unique association. The metabolic factor moderately correlated with gastrointestinal ($r_g$ = 0.39) and immune factors ($r_g$ = 0.25). The gastrointestinal disease factor had the most significant independent association with MDD (β = 0.63, $P = 3.04 \times 10^{-30}$). The gastrointestinal and immune disease factors had the highest correlation among the disease factors ($r_g$= 0.59). After accounting for the genetic overlap with the other three factors, the immune disease factor was no longer significantly associated with MDD (β = 0.08, $P$ = 0.156).

**1.5 Multiple Regression on Individual Traits.** A multiple regression model was used to analyse the individuals' traits that comprised the disease clusters with a significant independent association (Supplementary Figure 2; Supplementary Table 4). This was performed to identify traits with more substantial individual associations with MDD beyond the shared variance that may be driving the association. The regression model included 12 cardiovascular, metabolic, and gastrointestinal disease cluster traits. Two traits demonstrated a significant independent association, including irritable bowel syndrome ($\beta = 0.39$, $P = 4.73 \times 10^{-12}$) and peptic ulcer disease ($\beta = 0.28$, $P = 1.53 \times 10^{-03}$), two disorders within the gastrointestinal disease cluster. No trait showed a unique association with MDD in the cardiovascular or metabolic cluster. The variance of MDD explained by the individual traits was smaller than the disease factors themselves, with these individual traits explaining 43% of the genetic component of MDD.

### 2.0 Disease-MDD Latent Factor Sensitivity Analysis (SNP-based heritability Z > 4)

Latent factor analysis was performed using traits with a SNP-based heritability z score greater than four, calculated using LDSC. This threshold was used to compare against the alternative threshold as a sensitivity analysis and investigate trait selection.

We identified 27 traits that reached this threshold, with 20 significantly correlated with MDD ($\alpha < 1.85 \times 10^{-3}$; 0.05/number of traits). This threshold differed from a $h^2_{SNP}$ $Z > 7$ by the inclusion of peripheral artery disease (PAD; $h^2_{SNP}$ $Z = 6.70$; N = 483,078) in the cardiovascular cluster, iron deficiency anaemia (IDA; $h^2_{SNP}$ $Z = 5.18$; N = 480,941) in the metabolic cluster; Hashimoto's disease (HD; $h^2_{SNP}$ $Z = 5.18$; N = 395,640), and psoriasis vulgaris (PSV; $h^2_{SNP}$ $Z = 5.31$; N = 483,174) in the immune cluster (Supplementary Table 5). Models were created for each disease cluster (Supplementary Figure 3a-c). Each trait was loaded on a common latent factor, with MDD regressed onto that factor. Unit variance identification was used for model identification. See Supplementary Table 6 for full model results.

**2.1 Cardiovascular Disease Latent Factor.** This cardiovascular disease cluster included PAD in this model. The model fit was acceptable (CFI = 0.910; SRMR = 0.076). This group had the second largest standardised factor loadings of the four disease clusters (Mean $|\lambda| = 0.70$, Range $|\lambda|$ 0.93 – 0.38). The strongest loading was HF ($\lambda = 0.93$), followed by PAD ($\lambda = 0.80$), CAD ($\lambda = 0.73$), STK ($\lambda = 0.69$), and then AF ($\lambda = 0.38$). The cardiovascular disease factor was significantly associated with MDD ($\beta = 0.31$; $P = 8.50 \times 10^{-38}$). This factor explained approximately 9% of the genetic component of MDD.

**2.2 Metabolic Disease Latent Factor.** This latent factor model contained IDA as an additional metabolic trait. To amend a Heywood case, the MSYN variance was fixed to greater than 0.001, and its factor loading to less than 1. The model fitted well with a CFI = 0.969 and SRMR of 0.069. The metabolic disease cluster had the highest mean standardised factor loadings out of the four, although only marginally higher than the cardiovascular group (Mean |λ| = 0.71, Range |λ| 1 – 0.36). The factor loadings were as follows: T2D (λ = 0.70), MSYN (λ = 1), TG (λ = 0.78), HDL (λ = -0.72), IDA (λ = 0.36). The metabolic common factor was significantly associated with MDD (β = 0.23; *P* = 1.35 x $10^{-48}$). The residual variance of MDD was 0.95, with the latent factor model explaining 5% of the genetic component of MDD.

**2.3 Gastrointestinal Disease Latent Factor.** There was no difference in this cluster compared to using a SNP-based heritability z score greater than seven.

**2.4 Immune Disease Latent Factor.** The immune latent factor model contained two extra traits using this threshold: HD and PSV. Due to ASTH and AD's strong correlation, high residual variance, and previous reporting of an immune subgroup[1], these traits were fixed to correlate within the model. This improved model fit from a CFI of 0.644 and SRMR of 0.109 to a CFI of 0.960 and SRMR of 0.061. The cluster had the smallest standardised factor loadings of the four (Mean |λ| = 0.36, Range |λ| 0.65 – 0.20). The largest immune factor loading was HD (λ = 0.65), followed by PSV (λ = 0.49), ASTH (λ = 0.32), AD (λ = 0.25), IBD (λ = 0.25), and lastly, MS (λ = 0.20). This latent factor model was significantly associated with MDD (β = 0.66; *P* = 2.14 x $10^{-31}$) and explained a substantial portion of MDD genetic variance at 44%.

**2.5 Four-Factor Model.** The four individual models were added together with the parameters retained to identify the independent associations of each factor with MDD. MDD was regressed onto each factor in a multiple regression model, with each factor correlating (Supplementary Figure 4; Supplementary Table 7). The resulting model fit was below the threshold (CFI = 0.848; SRMR = 0.075).

The four-factor model captured 58% of the genetic component of MDD (calculated as 1 - 0.42), with the residual variance of MDD estimated at 0.42. Clusters demonstrating a significant independent association with MDD were metabolic (β = -0.14; *P* = 0.015) and immune (β = 0.57; *P* = 0.017). Cardiovascular (β = -0.07; *P* = 0.38) and gastrointestinal (β = 0.32; *P* = 0.82) clusters had no significant independent association. The cardiovascular factor showed strong correlations with the metabolic ($r_g$ = 0.59), gastrointestinal ($r_g$ = 0.41), and immune factors ($r_g$ = 0.59). The metabolic was moderately correlated with the other clusters (gastrointestinal: $r_g$ = 0.40; immune: $r_g$ = 0.51). The gastrointestinal and immune cluster had the strongest correlation ($r_g$= 0.77).

This threshold was not continued due to poor model fit and lesser-powered traits ($4 < Z < 7$) acting as influential outliers and disproportionately influencing correlations in modelling. Specifically, the immune cluster was heavily manipulated, reducing factor loadings of other well-powered traits and inflating the immune group's independent association with MDD. The latent factor loadings were skewed towards HD and PSV, distorting the factor loadings and emphasising the contribution of the weaker traits relative to the more reliable, well-powered immune traits. This is evident when compared to the $h^2_{SNP}$ $Z > 7$ models. Correlations between the immune and other factor groups are reduced, all factor loadings of the immune traits increase, and the independent association of the immune factor on MDD attenuates, enabling the independent associations of cardiovascular and gastrointestinal factors to emerge.

### 3.0 Multivariate GWAS.

**3.1 Second-order Latent Factor Models.** Second-order latent factor models were created for multivariate GWAS on the shared genetic variance between the disease clusters and MDD. A second-order latent factor (*disease*-MDD factor) was added to each disease latent factor model (Supplementary Figures 5a-d; Supplementary Table 8). The disease common factor and MDD loaded onto the second-order factor with their unstandardised factor loadings constrained to be equal, i.e., the factor loading from MDD and the disease latent factor are specified to have equal variance loading on the second-order factor. All four models demonstrated acceptable fit (CFI > 0.90, SRMR < 0.10).

The resulting estimated model parameters can describe the association of each variable (MDD and the disease cluster) with the downstream multivariate GWAS and each other. First, the variance of the second-order factors indicates the degree of shared variance between the disease latent factor and MDD, i.e., a larger captured variance suggests greater shared genetic architecture. Second, the resulting standardised factor loadings specify the proportion of variance accounted for by the disease-MDD latent factor. This indicates how much of the total variance of the variable is associated with the second-order factor and thus, its strength of association with the conducted multivariate GWAS. Contrarily, the residual variance of a variable demonstrates how much of the total variance is not explained by the second-order factor.

The cardiovascular-MDD latent factor had a standardised variance of 0.30, indicating a moderate overlap between MDD and the cardiovascular factor. This model had similar standardised loadings and residual variances between the cardiovascular factor ($\lambda = 0.56$; $\theta_{CVD} = 0.69$) and MDD ($\lambda = 0.55$; $\theta_{MDD} = 0.70$). This model demonstrates that a multivariate GWAS conducted on the second-order latent factor will explain a moderate portion of the cardiovascular latent factor and MDD variance.

Likewise, the metabolic-MDD model had near equivalent standardised loading and residual variances onto the higher-order factor for the metabolic factor ($\lambda = 0.49$; $\theta_{Metab} = 0.76$) and MDD ($\lambda = 0.50$; $\theta_{MDD} = 0.75$). The metabolic-MDD factor had a standardised variance of 0.25, demonstrating a relatively modest genetic variance shared between the metabolic cluster and MDD. As a result, a multivariate GWAS will reflect a modest association between these variables.

The gastrointestinal-MDD model captured the most variance ($\theta_{Gastro\text{-}MDD} = 0.54$) across the second-order factor models. This is in line with previous results, following the trend observed in LDSC and the individual models, with the gastrointestinal cluster having a strong association with MDD compared to the other models. The gastrointestinal-MDD model showed a larger standardised loading of the disease factor ($\lambda = 0.94$; $\theta_{Gastro} = 0.12$) compared to MDD ($\lambda = 0.73$; $\theta_{MDD} = 0.46$). These differing proportions reflect the smaller variance captured by the gastrointestinal factors, leading to a larger standardised proportion of disease cluster variance. This model suggests that a downstream multivariate GWAS will reflect the gastrointestinal factor well and represents the strongest association with MDD across the models.

The immune-MDD factor captured all the immune factor variance ($\lambda = 1$; $\theta_{Immune} = 0.005$) and had the smallest standardised MDD loading ($\lambda = 0.45$; $\theta_{MDD} = 0.79$). Like the gastrointestinal model, the larger standardised proportion reflects the smaller variance of the immune disease factor. The immune-MDD factor variance was 0.21, suggesting the least shared variance between the immune cluster and MDD. A multivariate GWAS will correspond to all the immune latent factor variance and be associated with a smaller proportion of MDD variance than the other clusters.

**3.2 $Q_{SNP}$ Heterogeneity Test.** SNPs were filtered out based on whether they demonstrated significant $Q_{SNP}$ heterogeneity. All SNPs that were in LD with a significant $Q_{SNP}$ were also removed, i.e., removing the putative $Q_{SNP}$ locus. This quality control step ensures that the follow-up analysis excludes SNPs showing significant heterogeneity. Supplementary Table 9 shows the number of SNPs in the GWAS and the independent loci identified before and after $Q_{SNP}$ removal. The percentage of $Q_{SNP}$ heterogeneity reflects the proportion of independent loci significantly associated with the disease-MDD factor that do not operate via the factor but are associated with MDD and the disease factor independently. Of the identified loci, the immune-MDD factor exhibited the highest $Q_{SNP}$ heterogeneity with 13.5%, followed by cardiovascular-MDD (12.69%), then the metabolic-MDD factor (6.12%). The gastrointestinal-MDD factor had no independent genome-wide significant loci with high Q heterogeneity. This suggests that most loci associated with the shared liability of disease groups and MDD operate through common mechanisms.

**3.3 Independent Loci.** To identify independent loci associated with the shared liability between disease clusters and MDD, the multivariate GWAS summary statistics for each second-order latent factor were uploaded to FUMA[2] ($r^2$ = 0.60; distance ≥ 250kb; Supplementary Tables 10-13). We identified 537 independent loci associated with the metabolic-MDD latent factor, the largest among the latent factors. This was followed by CVD-MDD (N = 172), gastrointestinal-MDD (N = 170), and immune-MDD (N = 140). A large proportion of disease-MDD loci were previously unidentified for their respective disease traits (Proportion = previously unidentified loci/total loci; CVD-MDD = 70%; gastrointestinal-MDD = 75%; immune-MDD = 77%), while the metabolic-MDD had the smallest proportion (metabolic-MDD = 23%). Contrarily, the metabolic-MDD factor had the highest proportion of loci previously unidentified with MDD (74%), while the other disease-MDD factors had smaller proportions (CVD-MDD = 44%; gastrointestinal-MDD = 28%; immune-MDD = 24%). This is likely due to the well-powered traits used in the metabolic disease factor, leading to a skewing of metabolic signal compared to MDD. Independent loci unique to each factor (i.e., not observed in the other second-order factors) may point to genetic drivers of MDD comorbidity for their respective disease clusters. CVD-MDD latent factor had 66 unique loci, metabolic-MDD had 417, gastrointestinal-MDD had 42, and immune-MDD had 34 loci.

### 4.0 Phenome-wide Association Studies

PheWASs were performed to identify diseases/traits associated with the independent loci of the disease-MDD factors (Supplementary Figures 7a-d, Supplementary Tables 14-17). This analysis would help uncover other associations for a broader understanding of these factors, functionally characterise the identified loci to provide biological insight, and validate the genetic underpinnings of the latent factors. The PheWAS associations were further partitioned into diseases/traits associated with loci previously unidentified to MDD and univariate disease traits and loci unique to the second-order factor.

**4.1 Cardiovascular-MDD PheWAS.** The primary categories most strongly linked to CVD-MDD loci included anthropometric measures, psychiatric, and cardiovascular traits. Notable trait groups were BMI, obesity, blood pressure, educational traits, immune cells, and lipid metabolism, which exhibited the highest counts. Additionally, loci that had not previously been associated with CVD traits showed large counts in education, MDD, depressive symptom categories and CVD-related associations. The loci previously unidentified to MDD demonstrated the strongest associations with BMI, blood pressure, obesity, and immune cells. Those unique to this category had the most pronounced associations with BMI, blood pressure, and immune cells. These associations support possible mechanistic links between MDD inflammation[3], metabolic dysregulation and CVD-MDD comorbidity.

**4.2 Metabolic-MDD PheWAS.** This factor had the highest association counts in metabolic, anthropometry and haematological umbrella categories. Trait groups involving BMI, lipid metabolism, obesity, diabetes, and blood measures/biomarkers were the primary groups linked to this factor. Loci previously unidentified with metabolic disease traits had strong associations with anthropometric measures, educational traits, and tobacco use. Anthropometric and metabolic categories had high numbers in loci previously unidentified in MDD. While not in the leading overarching categories, psychiatric disorders/traits had moderate numbers in MDD, neuroticism, and depressive symptoms. Loci unique to this factor had the highest counts in anthropometry, metabolic groups, haematological and educational traits. The PheWAS analysis revealed a high association with BMI, lipid metabolism, obesity and diabetes, which may support the link to lipid and carbohydrate metabolism association with depression risk[4].

**4.3 Gastrointestinal-MDD PheWAS.** Psychiatric, anthropometry, and *other* categories are strongly associated with the gastrointestinal-MDD factor. These can be broken down further into trait groups: BMI, educational traits, MDD, obesity and depressive symptoms. BMI, MDD, obesity, and neuroticism had the highest associations for loci unidentified to disease traits. For loci previously unidentified in MDD, the groups' BMI/obesity measures, educational traits, and gastrointestinal disorders had high counts. Trait group BMI, obesity, immune disorders, and tobacco use were strongly associated with loci unique to the gastrointestinal factor.

**4.4 Immune-MDD PheWAS.** Lastly, the top categories of the immune-MDD factor PheWAS were psychiatric, anthropometry, and *other*. More specifically, trait groups with the highest counts included BMI, obesity, educational traits, MDD, and immune cells. BMI, obesity, and MDD were highest in loci previously unidentified with the immune traits. Loci previously unidentified in MDD had the highest count in immune cells, BMI, and obesity trait groups. BMI, immune cells, and brain morphology demonstrated the strongest association with loci unique to this factor.

#### 5.0 Gastrointestinal Factor Follow-up Analysis.

**5.1 Functional vs Non-functional GORD.** The gastrointestinal disease cluster demonstrated a significant association with MDD. This was hypothesized to be due to the psychosomatic component relating to functional gastrointestinal disorders. Functional gastrointestinal disorders are chronic conditions not linked to underlying gut pathology but related to gastrointestinal symptoms (e.g., nausea, pain, bowel habits), which are exacerbated by stress or psychological comorbidities[5]. These disorders are more closely associated with MDD than non-functional gastrointestinal disorders. GORD is a common functional gastrointestinal disease and had the strongest factor loading among the gastrointestinal traits. A GWAS investigating GORD identified associated loci that are depression-driven (i.e., functional GORD) and those that are obesity-driven (i.e., non-functional GORD)[6]. We investigated whether the gastrointestinal-MDD factor was more strongly associated with functional or non-functional GORD.

First, we utilised the loci from the GWAS on functional and non-functional GORD and compared their effect sizes in the gastrointestinal-MDD second-order factor. Supplementary Figure 8 illustrates the difference in effect sizes between depression-driven (functional) and obesity-driven (non-functional) genetic variants, highlighting a significant difference between the two groups (P = 0.026). Functional GORD-derived loci demonstrated larger effect sizes. Second, we performed LDSC using functional and non-functional GORD to identify genetic correlations with MDD and other gastrointestinal traits (Supplementary Figure 9). MDD showed a higher genetic correlation with functional GORD ($r_g$ = 0.56; SE = 0.03; $P$ = 1.19 x $10^{-107}$) compared to non-functional ($r_g$ = 0.39; SE = 0.02; $P$ = 1.44 x $10^{-59}$). These results suggest that the loci captured by the second-order factor are more closely related to the functional subtype of GORD. The gut-brain axis is hypothesised to be the driving mechanism influencing the psychopathological symptoms of functional gastrointestinal disorders[7]. The captured variance of this latent factor likely reflects the gut-brain axis connection with these disorders and MDD.

**5.2 Estimating direct and shared genetic effects.** We next investigated the evidence for correlated horizontal pleiotropy, whereby genetic risk for both traits may act through the same pathways, as opposed to direct vertical pleiotropy, whereby genetic variants act through one trait to influence the other. Firstly, we modelled the bivariate distribution of genome-wide effects of both GWAS (MDD and gastrointestinal common factor) using the latent causal variable (LCV) model[8]. The approach leverages the mixed-fourth moment (co-kurtosis) of this bivariate effect size distribution to examine whether the effect of genetic liability of trait one on trait two is proportionally larger than that of trait two on trait one. LCV was deployed to estimate a parameter termed the posterior median genetic causality proportion (GCP). A positive GCP value implies partial genetic causality of trait one on trait two, and *vice versa* for negative values. GCP values near zero imply that any genetic correlation estimated between two traits may arise from horizontal rather than vertical pleiotropy. The recommended threshold to define partial genetic causality in either direction is a significantly non-zero |GCP| > 0.6[43]. We found nominal evidence for a significantly non-zero positive posterior median GCP value (GCP = 0.23, posterior SE = 0.09,

*P* = 0.01). This means that there is some evidence that the effect of genetic liability to MDD on the gastrointestinal common factor is proportionally larger than the reverse direction (gastrointestinal common factor → MDD). However, the GCP magnitude is not large enough to draw strong conclusions from the LCV model (|GCP| < 0.6).

To further explore the relationship between genetic liability to MDD and the gastrointestinal common factor, we utilised the Causal Analysis Using Summary Effect (CAUSE) model, as outlined extensively elsewhere[9]. Briefly, the CAUSE mixture model estimates parameters of "shared" (correlated horizontal pleiotropy) and "causal" (direct or vertical pleiotropic effects) between two traits. Using the Bayesian model comparison approach, expected log pointwise posterior density (ELPD), a model that fixes the causal term to zero (sharing only) can be compared to a model that allows the causal term to be a free parameter (causal versus sharing only). We constructed models in both directions, i.e., gastrointestinal common factor → MDD and MDD → gastrointestinal common factor. We tested three different beta prior distributions with asymmetric hyperparameters (necessary for the model to be identifiable) on the $q$ parameter in CAUSE, with $q$ the proportion of variants that act through correlated pleiotropy – q ~ Beta(1,2), q ~ Beta(1,10), and q ~ Beta(1,100)[9]. Whilst CAUSE uses q ~ Beta(1,10) as its default prior choice, we place the most value on the results using the q ~ Beta(1,2), given we assume that the proportion of variants exhibiting correlated pleiotropy will likely be large. Interestingly, we found evidence in both directions that the causal model fit better than the sharing alone (Supplementary Figure 10), with the MDD → gastrointestinal common factor having more statistically significant improvements in model fit once the causal term was allowed to freely vary relative to gastrointestinal common factor → MDD. This is largely in line with the evidence from the LCV model that the effect of MDD to the gastrointestinal common factor is larger than in the reverse direction. However, we note that CAUSE may be inflated under the scenario of high levels of correlated pleiotropy – which means these results should be interpreted cautiously. Further investigation is required to understand the true extent of these results and the biological mechanisms involved.

### 6.0 Biological Annotations

**6.1 Gene-set Analysis.** Gene set enrichment was assessed using exploratory GSA-MiXeR results for Gene Ontology (GO) terms and Anatomical Therapeutic Chemical (ATC) drug classification codes for each second-order latent factor GWAS summary statistic and MDD-only (Supplementary Tables 18-27).

The cardiovascular-MDD factor included 50 GO gene sets with an enrichment score (ES) exceeding one. Of these, 27 were not observed in the MDD-only analysis and comprised cardiovascular and neurological-related gene sets. For instance, triglyceride binding exhibited the highest ES (21.37 ± 7.34), while dopamine-associated gene sets also displayed significant enrichment (e.g., dopamine binding; ES = 16.78 ± 8.07). Regarding drug ATC codes, the broader Level 2 gene sets, propulsives (A03F) and anxiolytics (N05B), were enriched for this factor with anxiolytics not enriched in the gene-set analysis for MDD only. Additionally, ten lower Level 3 ATC codes showed enrichment, with eight related to the nervous system and five used in psychiatric treatment (N05BE, N05AG, N05AL, N05AC, N05AF). Thioxanthene derivatives (N05AF) and other anti-dementia drugs (N06DX) were not observed in the MDD-only analysis.

The metabolic-MDD second-order factor exhibited the highest gene-set enrichment among the factors. The gene sets enriched in the GO terms (N = 405) were predominantly linked to metabolic pathways, particularly cholesterol, HDL, and LDL metabolism. Only nine of the identified GO gene-sets were enriched in MDD alone. Some gene sets related to the nervous system were not observed in the MDD-only analysis, including neuron projection and maintenance (ES = 2.13 ± 0.56) and neuronal signal transduction (ES = 1.75 ± 0.41). The metabolic-MDD factor uniquely demonstrated enrichment in Level 1 ATC drug codes, associated with medications for diabetes (A10; ES = 2.07 ± 0.33) and anti-obesity preparations (A08; ES = 1.18 ± 0.20). ATC codes at Levels 2 and 3 encompassed medications related to obesity and diabetes, with gene sets relating to the treatment of psychiatric conditions, such as diphenylbutylpiperidine derivatives (N05AG; ES = 4.23 ± 1.49) and azaspirodecanedione derivatives (N05BE; ES = 2.88 ± 1.13). ATC codes for the nervous system not seen in MDD only included gene sets relating to dopamine, including dopaminergic agents (N04B; ES = 2.29 ± 0.39), centrally acting sympathomimetics (N06BA; ES 3.77 ± 0.94) and dopamine agonists (NO4BC; ES = 2.24 ± 0.58).

Next, the gastrointestinal-MDD latent factor included 43 GO gene sets, with 16 not seen in the MDD-only analysis. Of these 16, some were associated with the nervous system, such as dopamine binding (ES = 14.99 ± 6.97), regulation of dopamine uptake involved in synaptic transmission (ES = 11.05 ± 4.36), and cerebral cortex GABAergic interneuron development (ES = 13.23 ± 5.66). Similar to the cardiovascular-MDD factor, propulsives (A03F; ES = 7.78 ± 3.29) and anxiolytics (N05B; ES = 5.07 ± 2.01) were enriched at ATC code Level 2, with antirheumatic agents uniquely enriched (M01C; ES = 8.52 ± 2.96), a class of drugs used to treat inflammatory arthritides[10]. Level 3 ATC codes included eight related to the nervous system and two associated with improving movement in the gastrointestinal tract: propulsives (A03FA; ES = 7.78 ± 3.31) and peripheral opioid receptor antagonists (A06AH; ES = 4.46 ± 1.10). Many of these were observed in MDD-only, with thioxanthene derivatives and (N05AF) and other anti-dementia drugs (N06DX) not observed.

The immune-MDD second-order factor showed a similar pattern of gene-set enrichment to the gastrointestinal-MDD factor. There were 34 GO gene sets enriched with the immune-MDD second-order factor, with 26 of these also enriched in the gastrointestinal-MDD factor. Many of these gene sets were linked to the nervous system. Following this trend, propulsives (A03F; ES = 8.81 ± 3.81) and anxiolytics (N05B; ES = 5.48 ± 2.28) Level 2 ATC codes exhibited enrichment, and all Level 3 ATC codes enriched in the immune-MDD factor were also observed in the gastrointestinal-MDD factor.

Next, we compared these results with MAGMA gene-set analysis. GSA-MiXeR does not produce formal P values; therefore, for a conservative approach, MAGMA can prefilter gene sets using Bonferroni correction (Supplement Tables 28-29). For GO gene sets, we observed four that were significantly associated with the CVD-MDD factor. Of these, three were observed in MDD-only, with gene set commissural neuron axon guidance ($P = 4.28 \times 10^{-6}$) not observed. The metabolic-MDD factor demonstrated 17 significant GO gene-sets in MAGMA analysis, with 16 not seen in MDD-only. Three gene-sets also demonstrated high enrichment in GSA-MiXeR (ES > 10) and were related to lipid and cholesterol transport. Seven gene-sets were significant for the gastrointestinal-MDD factor, with three not seen in MDD-only. These three gene sets were associated with the nervous system, including branching morphogenesis of a nerve, commissural neuron axon guidance, and somatodendritic compartment. There were no statistically significant GO gene sets for the immune-MDD factor. For the MAGMA ATC code analysis, only the MDD-only analysis and the immune-MDD factor observed significant gene sets. The ATC code G03EA (androgens and estrogens; MAGMA $P = 1.09 \times 10^{-4}$; ES = 3 ± 0.80) was significant in the immune-MDD factor which was also seen in MDD-only.

Compared to exploratory GSA-MiXeR analysis, the MAGMA saw a significant reduction in gene-set enrichment. A limitation of MAGMA is that the statistical significance relies on the GWAS sample size and the size of the gene set. Conversely, MiXeR-GSA has been shown to identify smaller gene sets with potentially higher biological relevance[11]. Our analysis accentuates this difference, with MAGMA demonstrating significance with larger gene sets compared to GSA-MiXeR, particularly with the ATC code analysis, which has relatively smaller gene sets.

**6.2 Transcriptome-wide Structural Equation Modelling.** Transcriptome-wide structural equation modelling was performed to identify genes associated with second-order latent factors via imputed tissue gene expression. This approach integrates transcriptome-wide association studies, a method that uses expression quantitative trait loci (eQTLs) data to identify genes whose expression is associated with each trait, with genomic SEM, which identifies gene expression patterns in a model to determine gene associations with the second-order latent factor. Additional analyses included the aggregated Cauchy association test (ACAT) and colocalisation (coloc). This helped identify genes that had a significant effect across tissues by meta-analysing p-values (ACAT hits) and determine that a variant has a high likelihood (PP4 > 0.80) of both being associated with a second-order factor and changes in gene expression, suggesting a gene may be causally involved (coloc genes) (Supplementary Tables 30-34). A TWAS was also performed for univariate MDD-only, with ACAT and colocalisation, for comparison with the second-order factors. Each tissue's mean proportion of colocalised genes (coloc genes/total genes in tissue) was normalised to z scores to investigate tissues with a high proportion of PP4 signal (z score > 1.96) and compare across disease-MDD factors (Supplementary Tables 35-39).

The CVD-MDD latent factor had 877 significant genes across tissues (P < 0.05/number of genes per tissue). Gene p-values were meta-analysed over the tissues expressed, determining 497 genes with a significant effect (ACAT hits). From these genes, 350 were not observed in MDD, and 221 were not in the other second-order factors. Of the ACAT hits, the PP4 colocalisation model was strongest for 179, i.e., an underlying variant was associated with the second-order factor and changes in gene expression. This second-order factor saw high coloc signal in vagina (Z = 2.09), small intestine (Z = 2.46), and the adrenal gland tissue (Z = 2.14). This differed from MDD-only which did not have high signal in vagina and the adrenal gland tissue.

Next, 2107 unique genes were identified to be associated with the metabolic-MDD factor. These were prioritised to genes with a significant effect across tissues using ACAT (N = 1959). These genes were largely unidentified in MDD-only analysis (N = 1815). This factor had the lowest proportion of coloc PP4 signal across ACAT hits, with 25% (418/1959 ACAT genes). Stomach (Z = 2.46), colon (Z = 2.33), and brain spinal cord tissue (Z = 2.43), demonstrated a high proportion of coloc genes. None of these tissues were observed in the MDD-only analysis.

We identified 690 genes associated with the gastrointestinal-MDD factor, with 406 having a significant effect across tissues (ACAT hits). Of the ACAT hits, 179 were unidentified in MDD-only analysis and 88 were unique to this factor. Colocalisation analysis reported that 122 of the ACAT hits preferred the PP4 model. Colon (Z = 2.18) and small intestine tissue (Z = 2.61) saw high coloc signal for this factor, with the colon tissue not detected in MDD-only.

Lastly, the immune-MDD factor had 670 genes associated with 477 ACAT hits. This factor had many genes not identified in the MDD-only analysis (N = 300). From the ACAT hits, 189 were not observed in the other factors. The PP4 colocalisation model was also strongest for 34% of ACAT hits. Like the gastrointestinal-MDD factor, colon (Z = 2.48) and small intestine tissue (Z = 2.53) demonstrated high coloc signal.

**6.3 gsMap.** This method was employed to identify cell types and cell-type-specific genes within the human brain cortex tissue linked with each second-order latent factor and MDD only. Two distinct cell type associations were noted: one between microglial cell type 2 and the immune-MDD factor and another between endothelial cells and the metabolic-MDD factor. All inhibitory cell types, astrocytes type 2, oligodendrocyte precursor cells, and layers 2/3, 4, and 6 of the brain cortices exhibited significant associations with all factors and MDD-only (Supplementary Tables 40-44).

The CVD-MDD factor demonstrated 34 cell-type-specific gene associations. Of these, 21 were not observed in MDD-only, and 19 were not associated with any other factor (Supplementary Table 45). This factor had the highest unique gene associations (N = 18; not observed in MDD-only or other second-order factors). Layer 6 of the brain cortex had the highest gene association count (N = 11), while this factor was the only second-order factor with gene associations in the inhibitory cell type 3 (PP1R13B, CDH9).

Next, we identified 28 cell-type-specific gene associations for the metabolic-MDD factor. A subset was not observed in MDD (N = 21) or the other factors (N = 17), with 16 genes unique to this factor (Supplement Table 46). The metabolic-MDD factor demonstrated these unique associations mostly within layer 6 (N = 8) and layer 2/3 (N = 6) of the brain cortex.

The gastrointestinal-MDD factor had 35 cell-type specific gene associations (Supplement Table 47). Among these, 13 were not observed in MDD, and only one gene was not found in the other factors. There were no unique gene associations for the gastrointestinal-MDD factor due to a high level of overlap with the immune-MDD factor, which shares 34 out of the 35 genes. Layer 2/3 and inhibitory cell type 2 had the highest gene association count (N = 9).

Finally, we observed 35 gene associations with the immune-MDD factor (Supplement Table 48). Fourteen were not seen in the MDD-only analysis, and one gene association was unique to this factor. This factor had one unique gene association, DUSP6, in layer six of the cortex. Layer 2/3 of the brain cortex had the highest gene association count (N = 9).

### Supplementary Figures

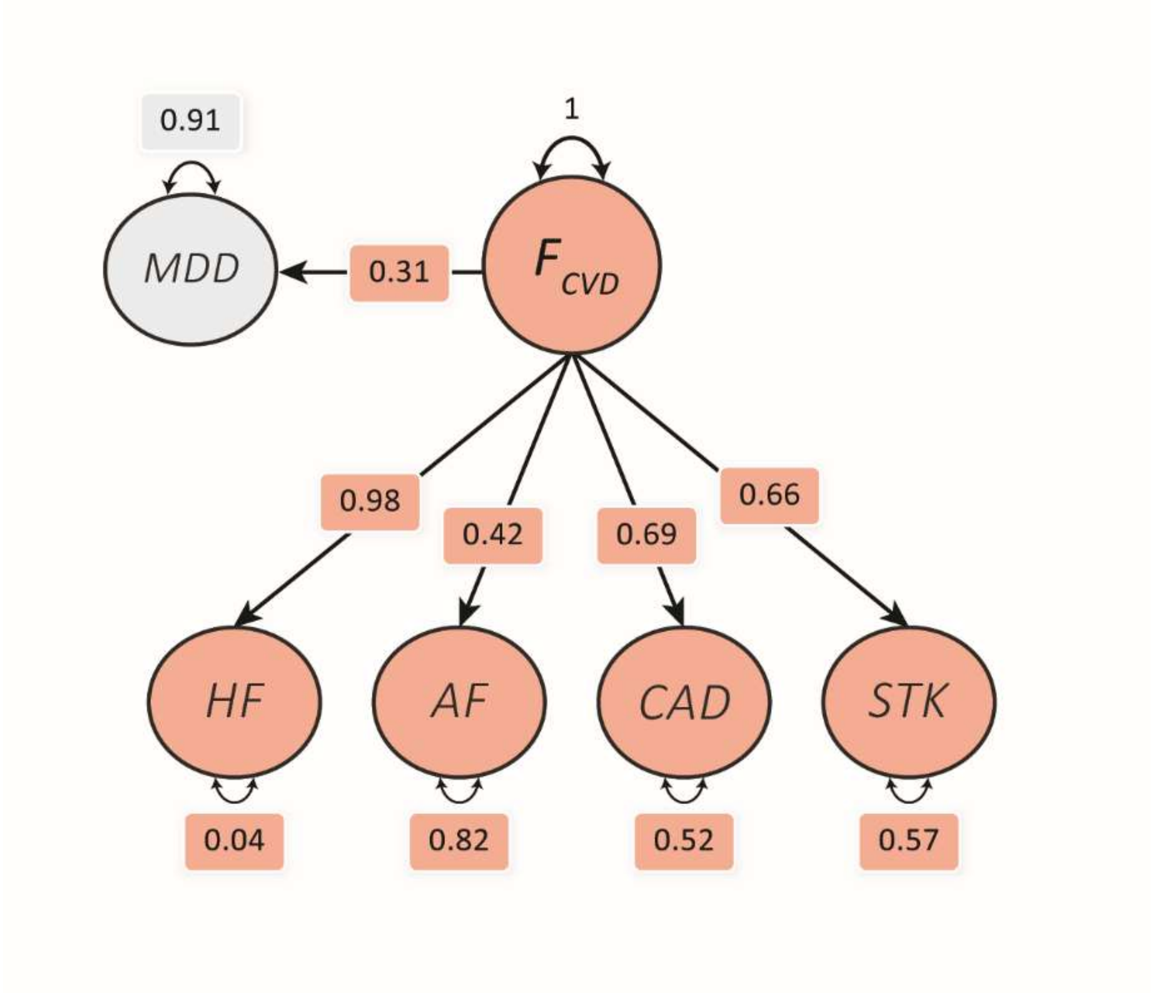

**Supplementary Figure 1a. Common factor model for cardiovascular traits and their combined association with Major Depressive Disorder (MDD).** The cardiovascular latent factor ($F_{CVD}$) captures the shared genetic variance across the cardiovascular traits with MDD regressed onto the factor. White outlines represent standardised estimated parameters. Acronyms: Atrial Fibrillation (AF), Coronary Artery Disease (CAD), Heart Failure (HF), Stroke (STK).

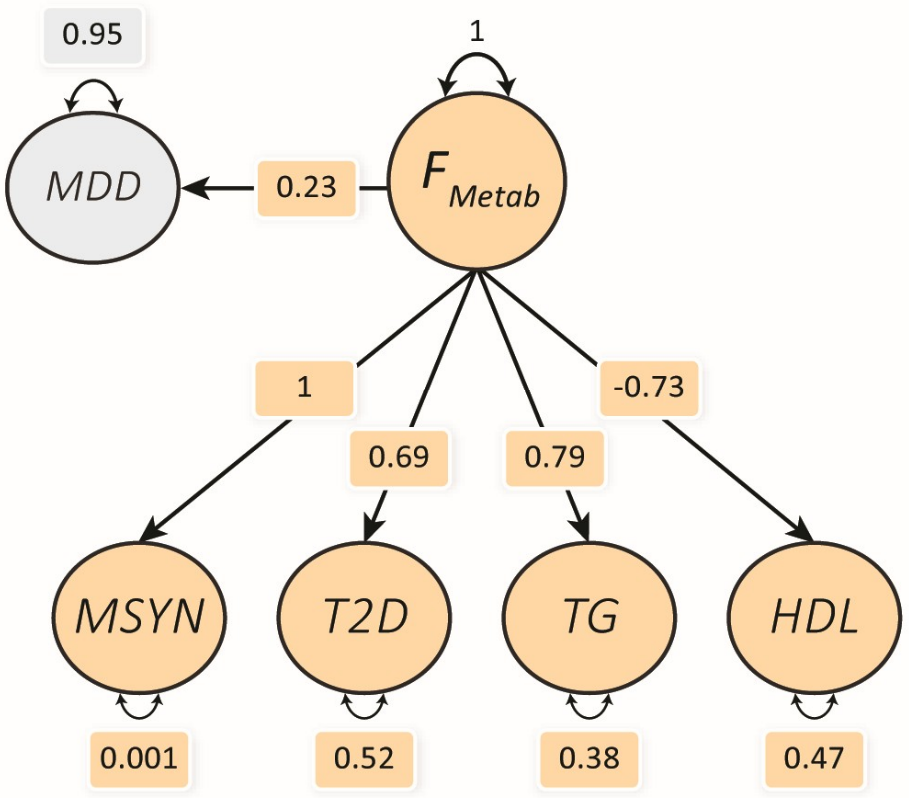

**Supplementary Figure 1b. Common factor model for metabolic traits and their combined association with Major Depressive Disorder (MDD).** The metabolic latent factor ($F_{Metab}$) captures the shared genetic variance across the metabolic traits with MDD regressed onto the factor. White outlines represent standardised estimated parameters. Acronyms: High-density lipoprotein cholesterol (HDL), Metabolic Syndrome (MSYN), Type 2 Diabetes (T2D), Triglycerides (TG).

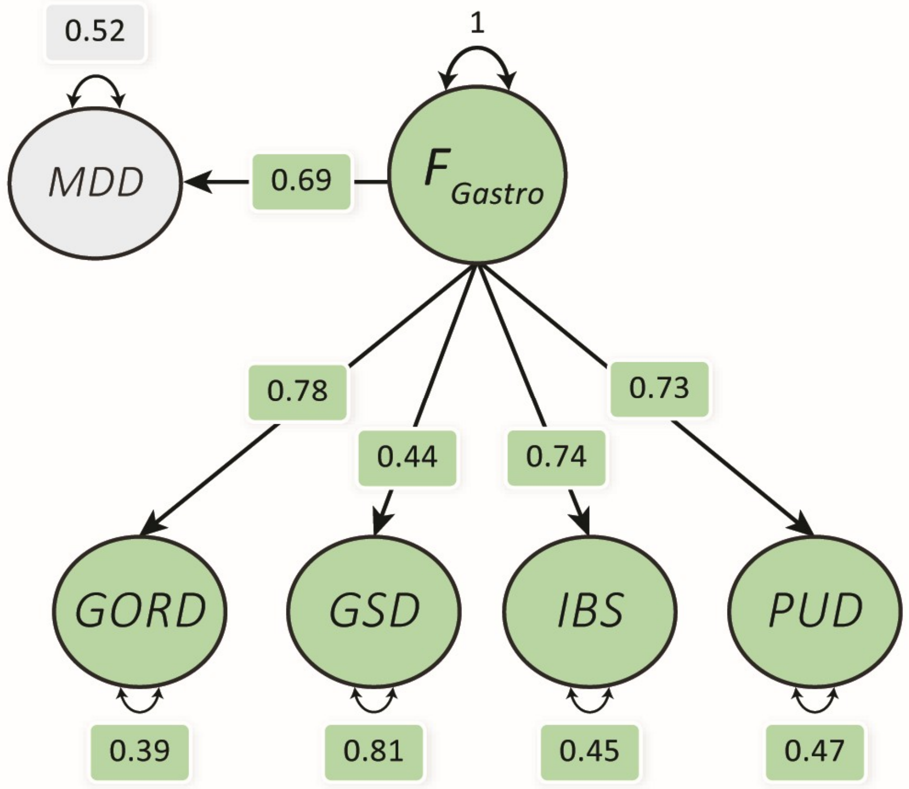

**Supplementary Figure 1c. Common factor model for gastrointestinal traits and their combined association with Major Depressive Disorder (MDD).** The gastrointestinal latent factor ($F_{Gastro}$) captures the shared genetic variance across the gastrointestinal traits with MDD regressed onto the factor. White outlines represent standardised estimated parameters. Acronyms: Gastro-oesophageal Reflux Disease (GORD), Gallstone Disease (GSD), Irritable Bowel Syndrome (IBS), Peptic Ulcer Disease (PUD).

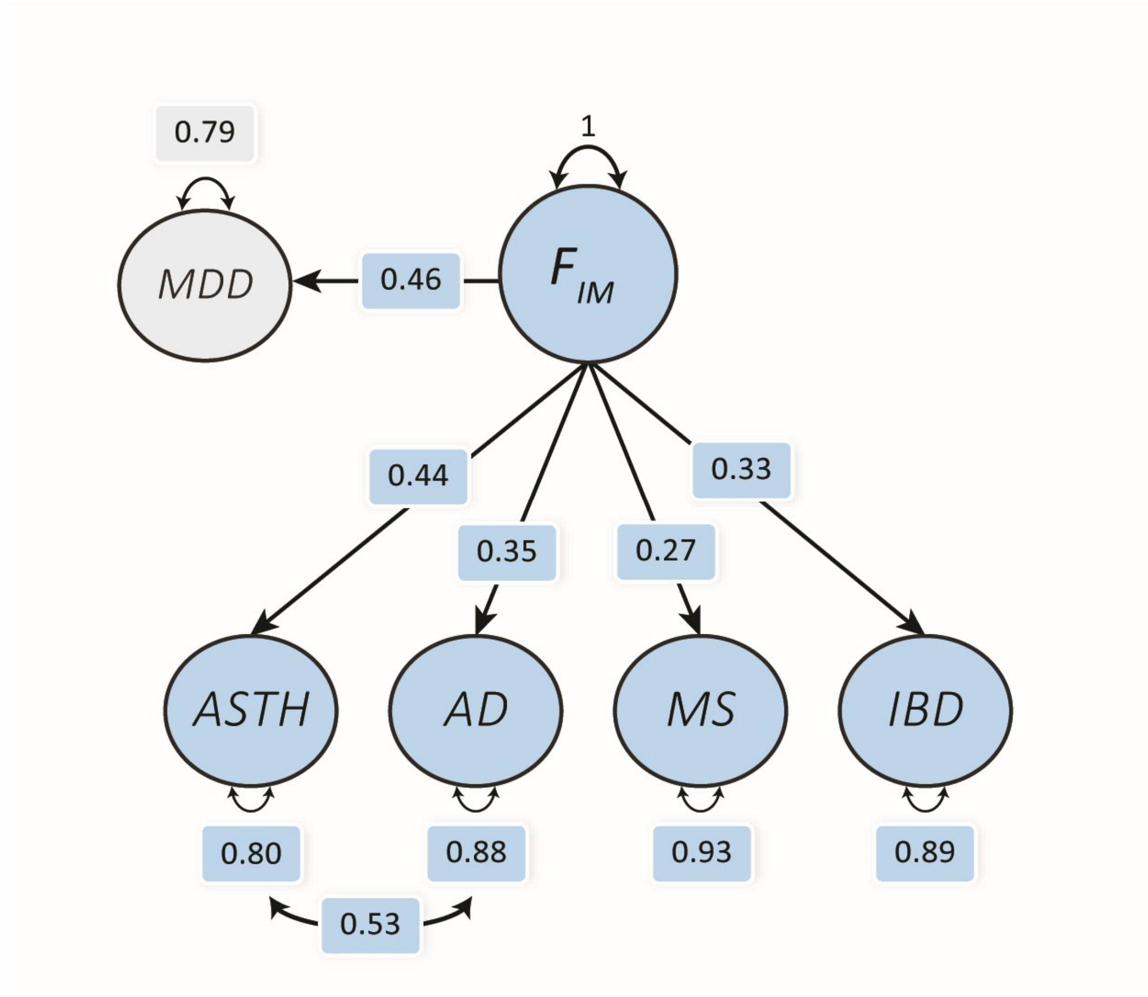

**Supplementary Figure 1d. Common factor model for immune traits and their combined association with Major Depressive Disorder (MDD).** The immune latent factor ($F_{IM}$) captures the shared genetic variance across the immune traits with MDD regressed onto the factor. White outlines represent standardised estimated parameters. Acronyms: Atopic Dermatitis (AD), Asthma (ASTH), Inflammatory Bowel Disease (IBD), Multiple Sclerosis (MS).

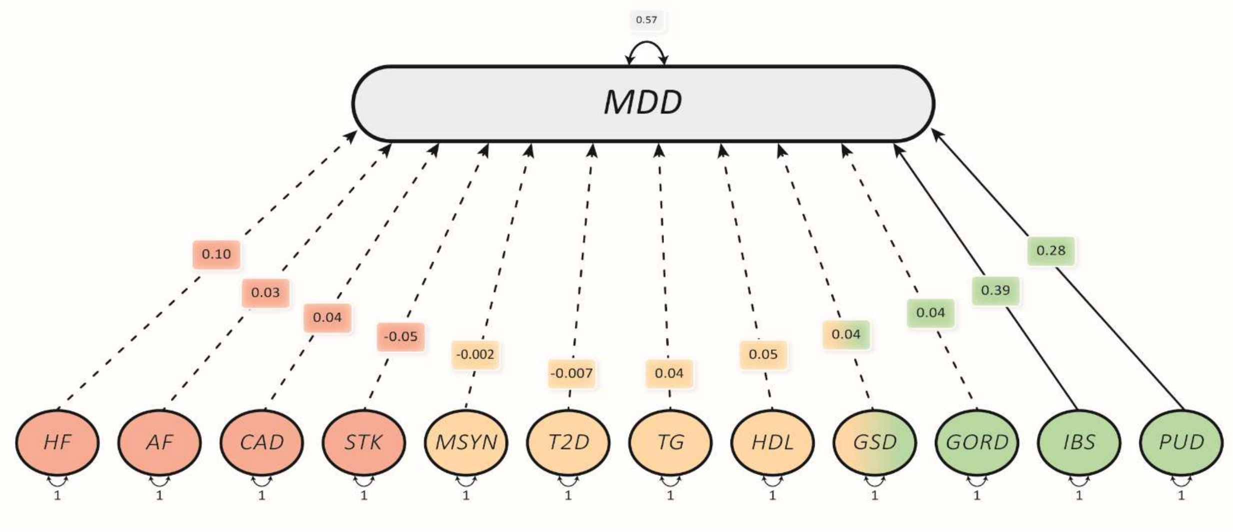

**Supplementary Figure 2. Multiple-regression model of the traits that make up the significant latent factor groups and their independent association on MDD.** A solid line demonstrates a significant independent association, while a dotted line indicates an insignificant association. White outlines represent standardised estimated parameters, with all traits correlating with each other. Acronyms: Atrial Fibrillation (AF), Coronary Artery Disease (CAD), Heart Failure (HF), Stroke (STK), High-density lipoprotein cholesterol (HDL), Metabolic Syndrome (MSYN), Type 2 Diabetes (T2D), Triglycerides (TG), Gastro-oesophageal Reflux Disease (GORD), Gallstone Disease (GSD), Irritable Bowel Syndrome (IBS), Peptic Ulcer Disease (PUD).

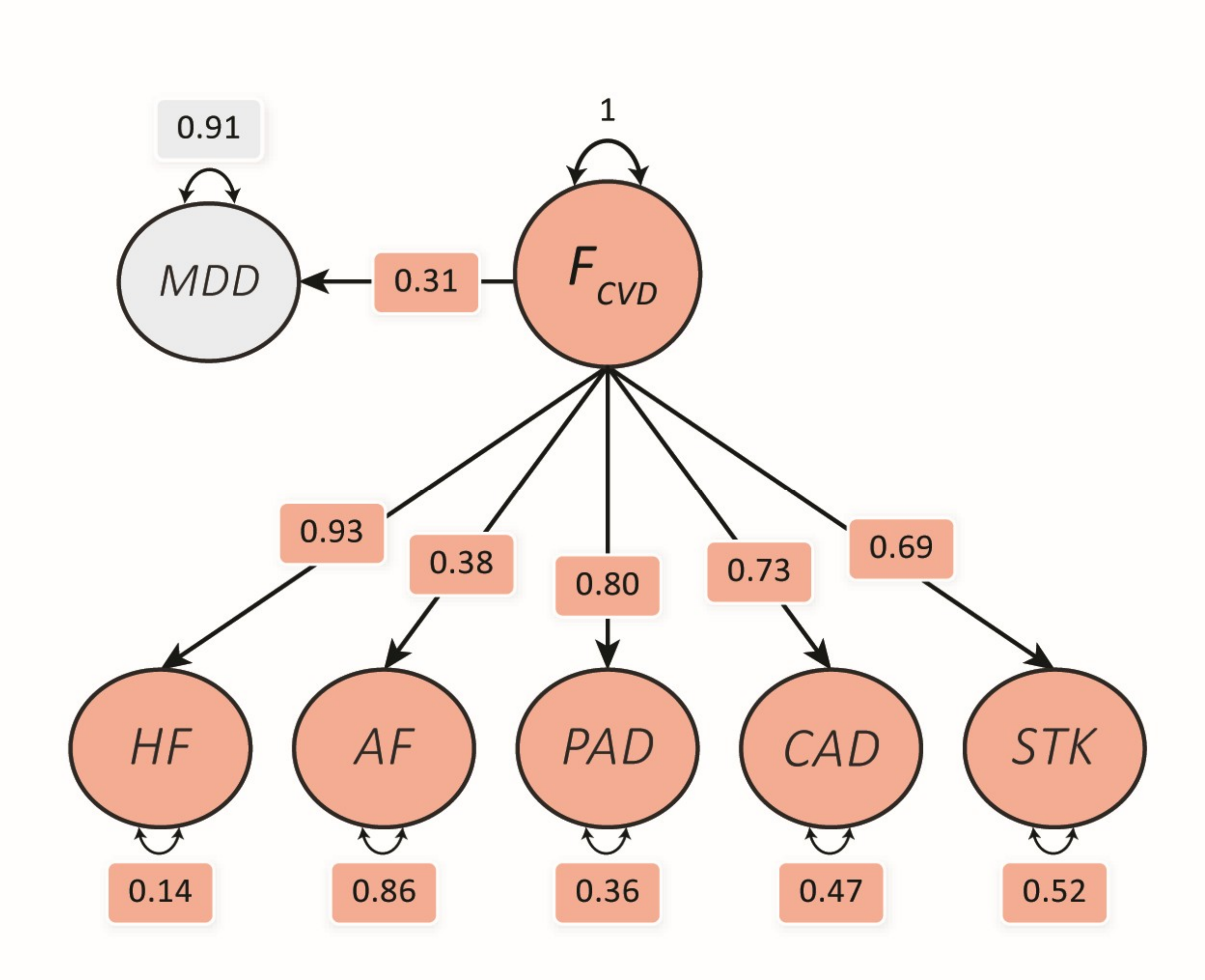

**Supplementary Figure 3a. Common factor model for cardiovascular traits and their combined association with Major Depressive Disorder (MDD) using $h^2_{SNP}$ Z > 4.** The cardiovascular latent factor ($F_{CVD}$) captures the shared genetic variance across the cardiovascular traits with MDD regressed onto the factor. White outlines represent standardised estimated parameters. Acronyms: Atrial Fibrillation (AF), Coronary Artery Disease (CAD), Heart Failure (HF), Stroke (STK), Periphery Artery Disease (PAD).

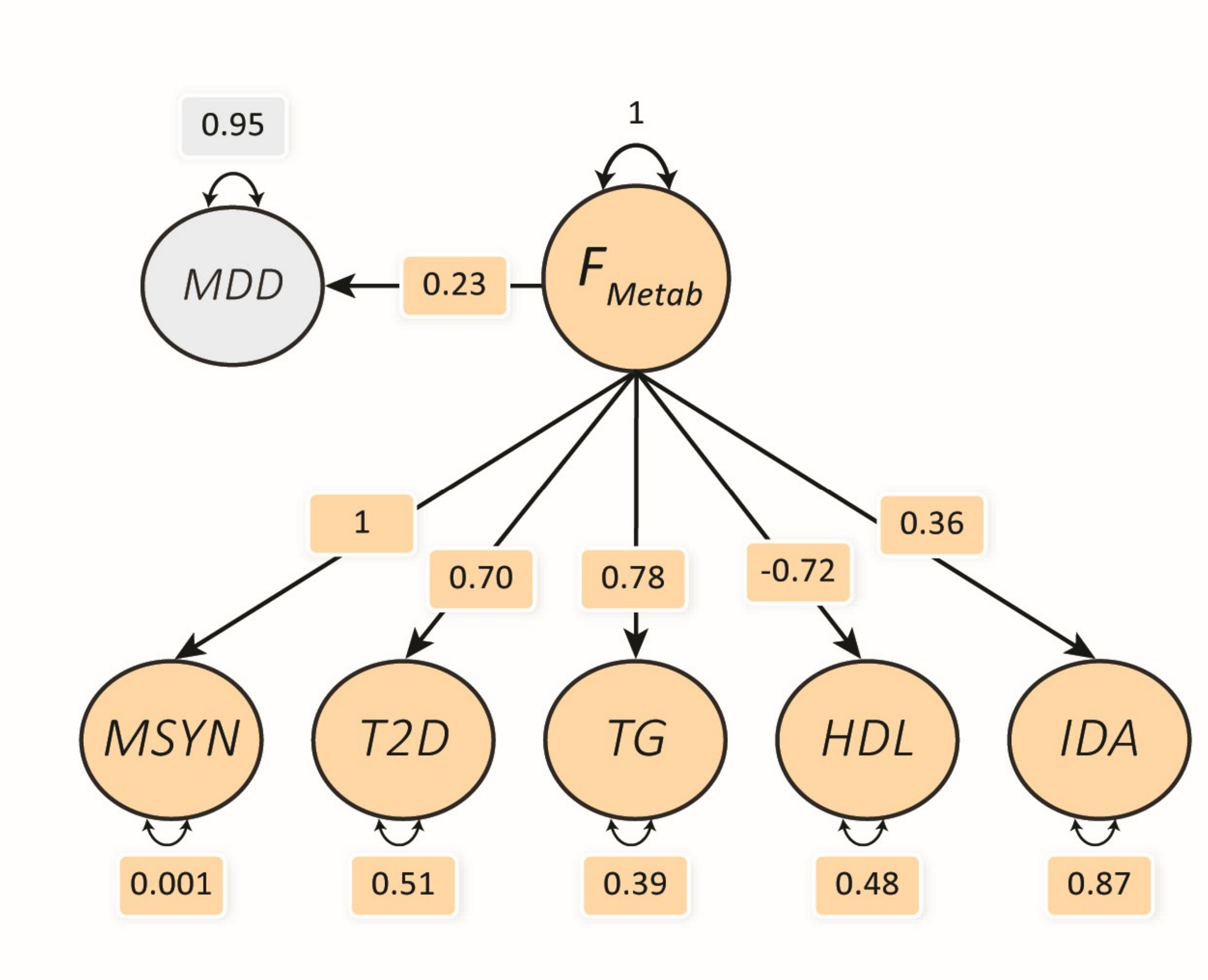

**Supplementary Figure 3b. Common factor model for metabolic traits and their combined association with Major Depressive Disorder (MDD) using $h^2_{SNP}$ Z > 4.** The metabolic latent factor ($F_{Metab}$) captures the shared genetic variance across the metabolic traits with MDD regressed onto the factor. White outlines represent standardised estimated parameters. Acronyms: High-density lipoprotein cholesterol (HDL), Metabolic Syndrome (MSYN), Type 2 Diabetes (T2D), Triglycerides (TG), Iron Deficiency Anemia (IDA).

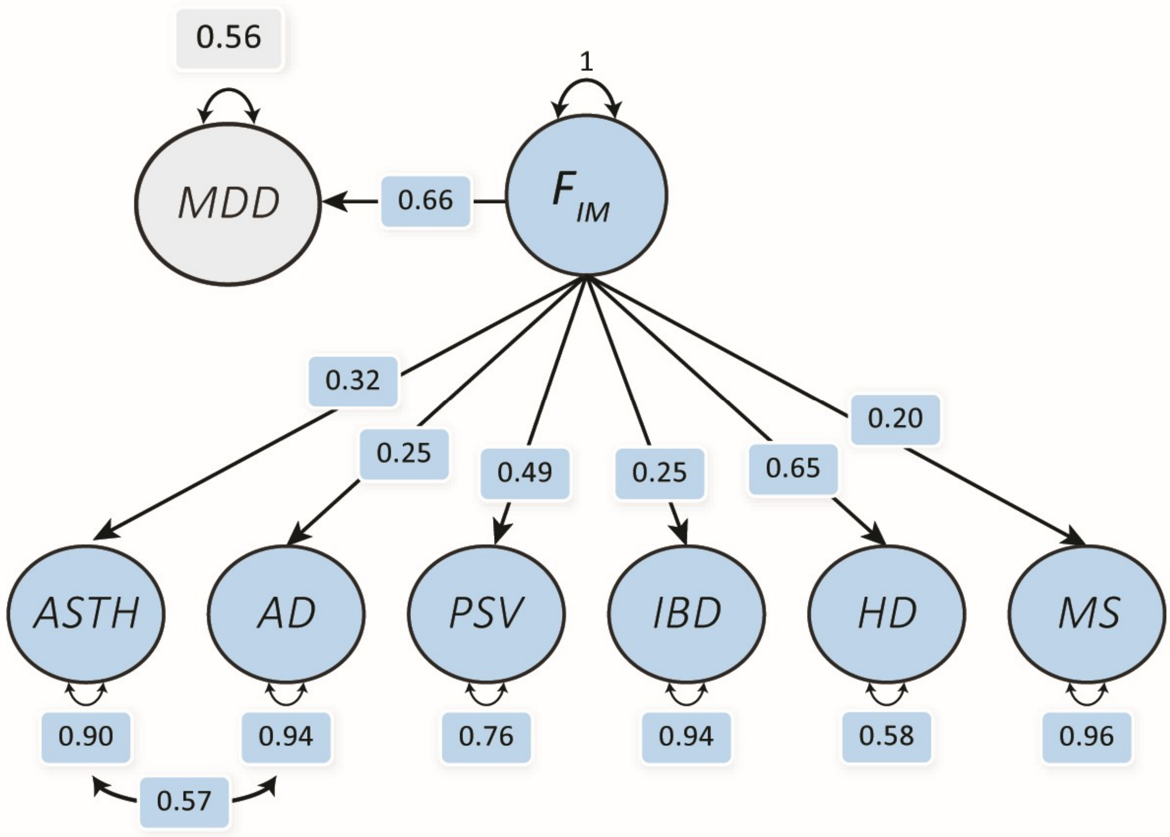

**Supplementary Figure 3c. Common factor model for immune traits and their combined association with Major Depressive Disorder (MDD) using $h^2_{SNP}$ Z > 4.** The immune latent factor ($F_{IM}$) captures the shared genetic variance across the immune traits with MDD regressed onto the factor. White outlines represent standardised estimated parameters. Acronyms: Atopic Dermatitis (AD), Asthma (ASTH), Inflammatory Bowel Disease (IBD), Multiple Sclerosis (MS), Psoriasis (PSV), Hashimoto's Disease (HD).

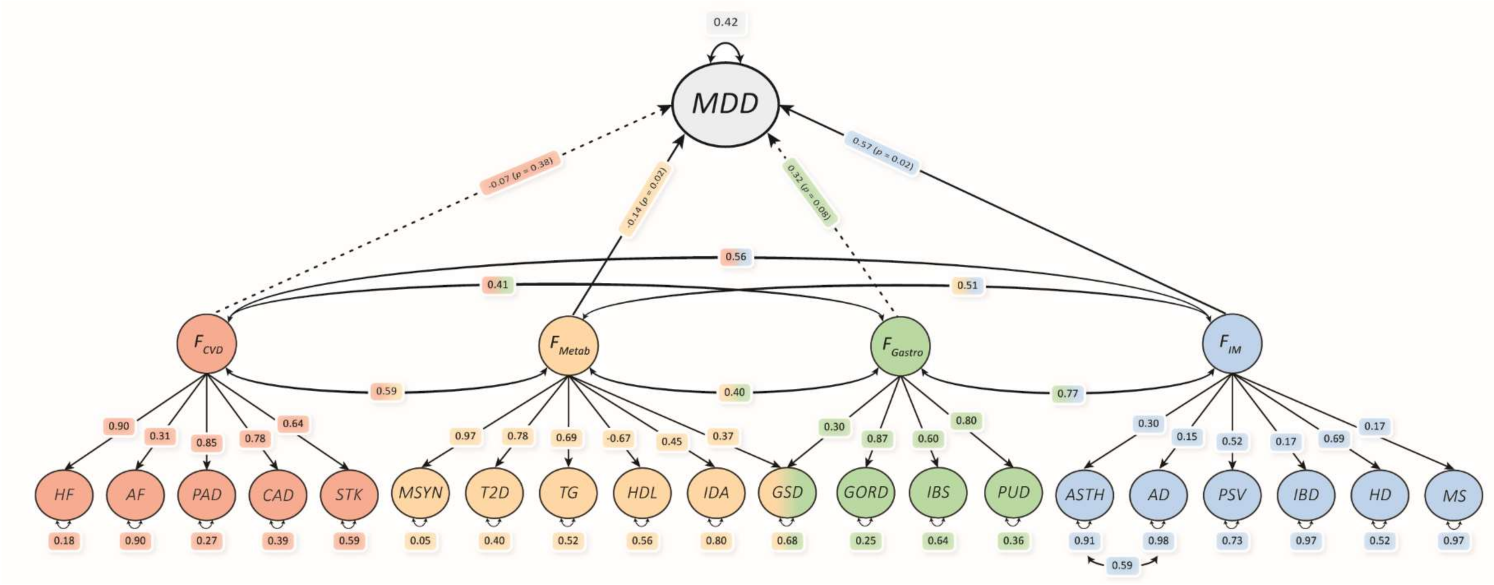

**Supplementary Figure 4. Multiple regression model of disease group latent factors (F) and their independent association with major depressive disorder (MDD) estimate using genomic structural equation modelling using $h^2_{SNP}$ Z > 4.** A solid line demonstrates a significant independent association, while a dotted line indicates an insignificant association. White outlines represent standardised estimated parameters. Acronyms: Cardiovascular (CVD), Metabolic (Metab), Gastrointestinal (Gastro), Immune (IM).

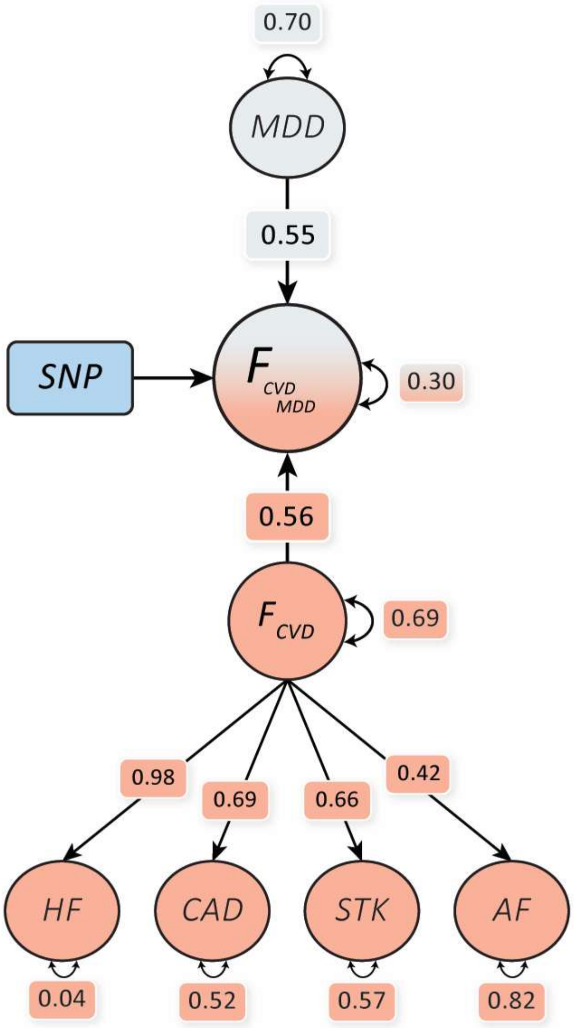

**Supplementary Figure 5a. Cardiovascular (CVD)-Major Depressive Disorder (MDD) second-order latent factor model used for multivariate genome-wide association study estimated using genomic structural equation modelling.** The model estimates a SNPs association with the second order factor. Both MDD and the disease factor are constrained to contribute equal unstandardised factor loadings. White outlines represent estimated parameters. The figure demonstrates standardised model metrics. Acronyms: Heart Failure (HF), Coronary Artery Disease (CAD), Stroke (STK), Atrial Fibrillation (AF).

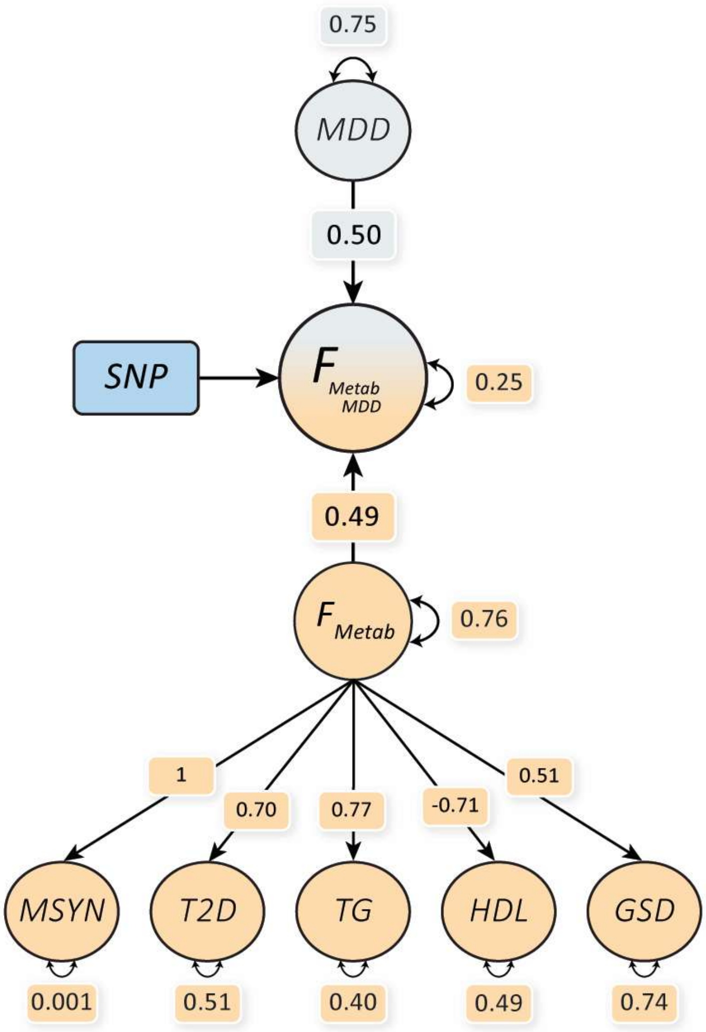

**Supplementary Figure 5b. Metabolic (Metab)-Major Depressive Disorder (MDD) second-order latent factor model used for multivariate genome-wide association study estimated using genomic structural equation modelling.** The model estimates a SNPs association with the second order factor. Both MDD and the disease factor are constrained to contribute equal unstandardised factor loadings. White outlines represent estimated parameters. The figure demonstrates standardised model metrics. Acronyms: Metabolic Syndrome (MSYN), Type 2 Diabetes (T2D), Triglycerides (TG), High-density lipoprotein cholesterol (HDL), Gallstone Disease (GSD).

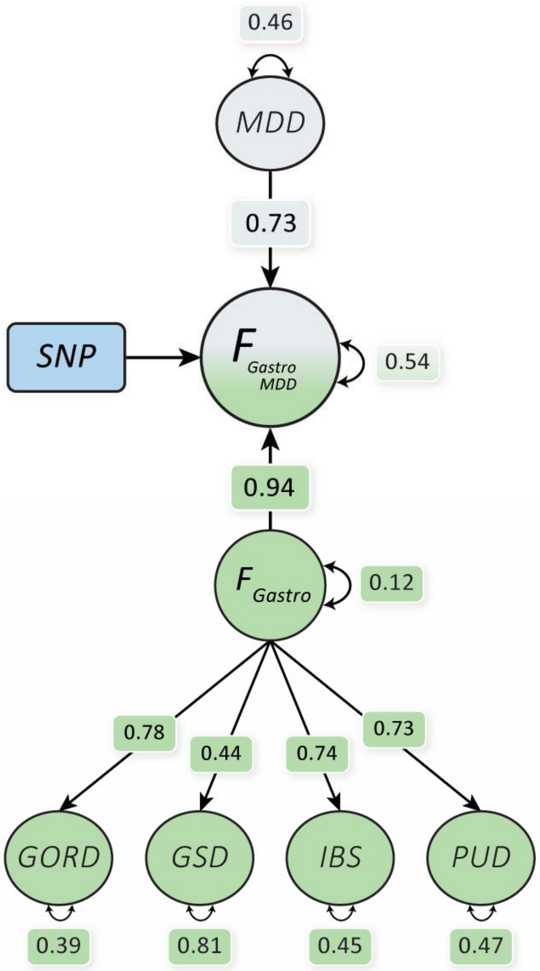

**Supplementary Figure 5c. Gastrointestinal (Gastro)-Major Depressive Disorder (MDD) second-order latent factor model used for multivariate genome-wide association study estimated using genomic structural equation modelling.** The model estimates a SNPs association with the second order factor. Both MDD and the disease factor are constrained to contribute equal unstandardised factor loadings. White outlines represent estimated parameters. The figure demonstrates standardised model metrics. Acronyms: Gastro-oesophageal Reflux Disease (GORD), Gallstone Disease (GSD), Irritable Bowel Syndrome (IBS), Peptic Ulcer Disease (PUD).

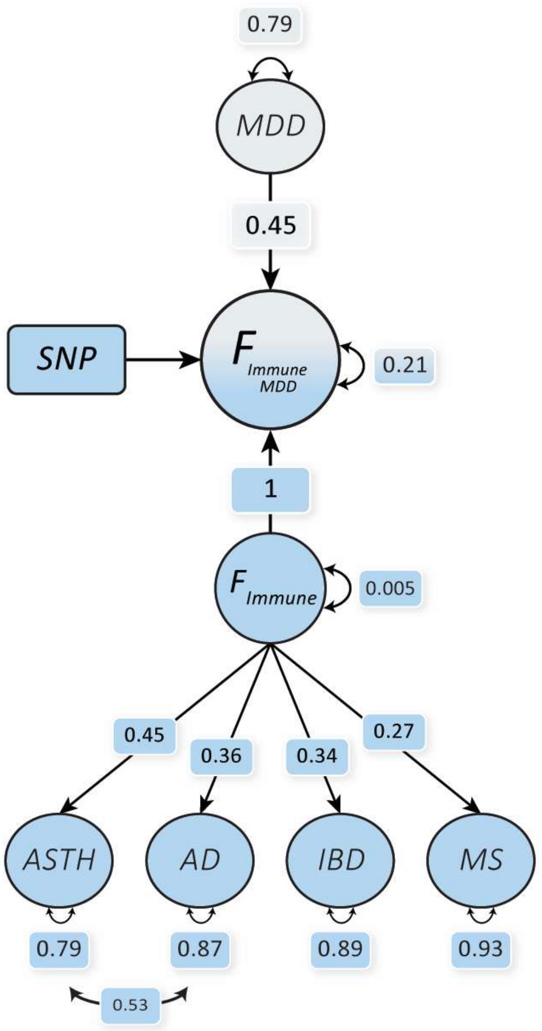

**Supplementary Figure 5d. Immune-Major Depressive Disorder (MDD) second-order latent factor model used for multivariate genome-wide association study estimated using genomic structural equation modelling.** The model estimates a SNPs association with the second order factor. Both MDD and the disease factor are constrained to contribute equal unstandardised factor loadings. White outlines represent estimated parameters. The figure demonstrates standardised model metrics. Acronyms: Asthma (ASTH), Inflammatory Bowel Disease (IBD), Atopic Dermatitis (AD), Multiple Sclerosis (MS).

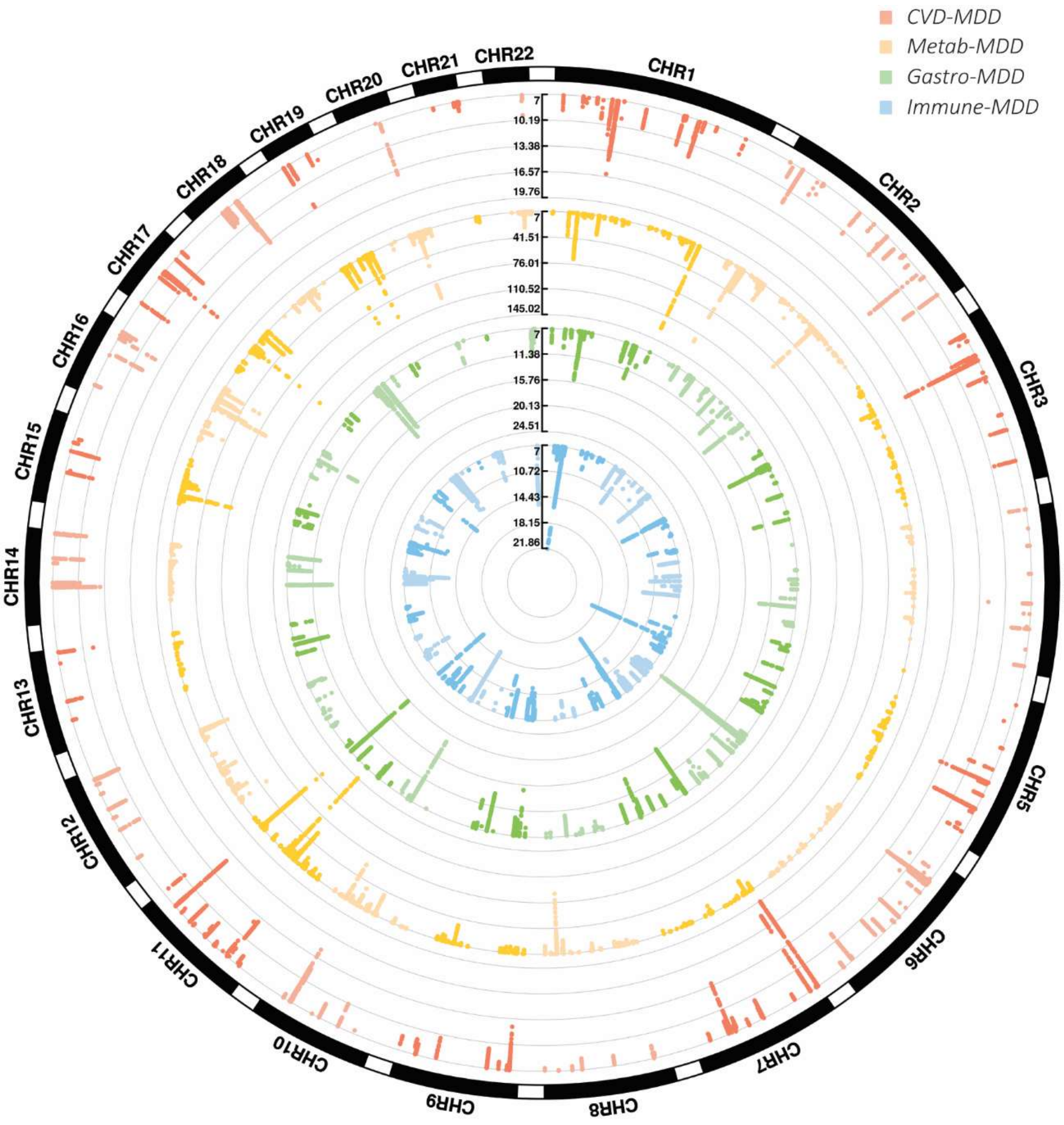

**Supplementary Figure 6. Circular Manhattan plot of genome-wide significant ($P < 5 \times 10^{-8}$) variants associated with the second-order factor disease models.** The multivariate genome-wide association study (GWAS) on second-order disease-major depressive disorder (MDD) latent factors was estimated using genomic structural equation modelling. The scale follows a negative log P value. Acronyms: Cardiovascular (CVD), Metabolic (Metab), Gastrointestinal (Gastro).

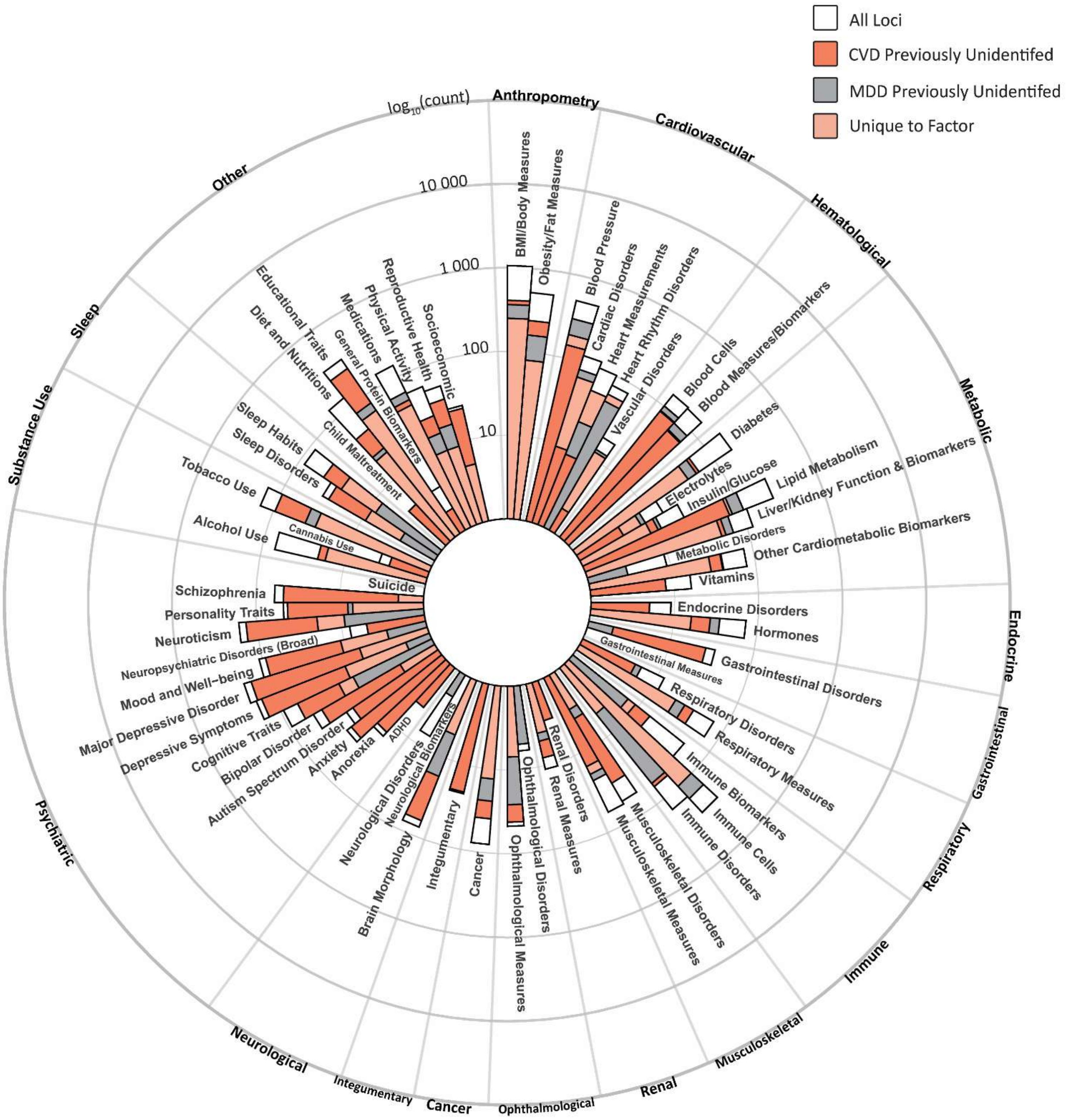

**Supplementary Figure 7a. Phenome-wide association studies of CVD-MDD latent factor, linking known disease/trait associations to identified independent loci.** Circular bar plots illustrate the number of associations per trait group on a logarithmic scale. The count of associations is further partitioned into loci previously unidentified to disease traits used in the multivariate GWAS, loci previously unidentified to major depressive disorder (MDD), and loci that are unique to each factor (i.e., loci are not associated with the other disease factors).

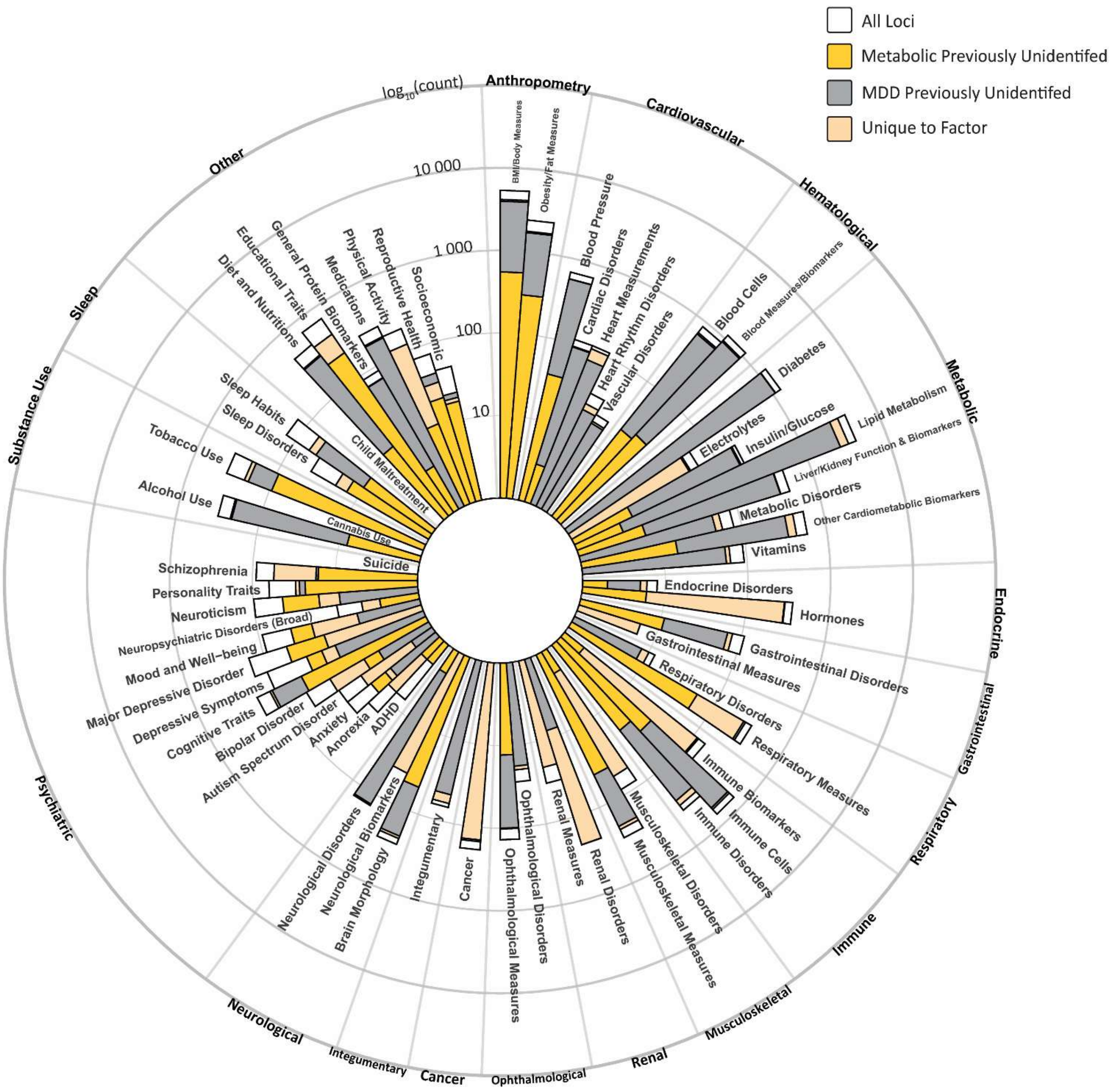

**Supplementary Figure 7b. Phenome-wide association studies of Metabolic-MDD latent factor, linking known disease/trait associations to identified independent loci.** Circular bar plots illustrate the number of associations per trait group on a logarithmic scale. The count of associations is further partitioned into loci previously unidentified to disease traits used in the multivariate GWAS, loci previously unidentified to major depressive disorder (MDD), and loci that are unique to each factor (i.e., loci are not associated with the other disease factors).

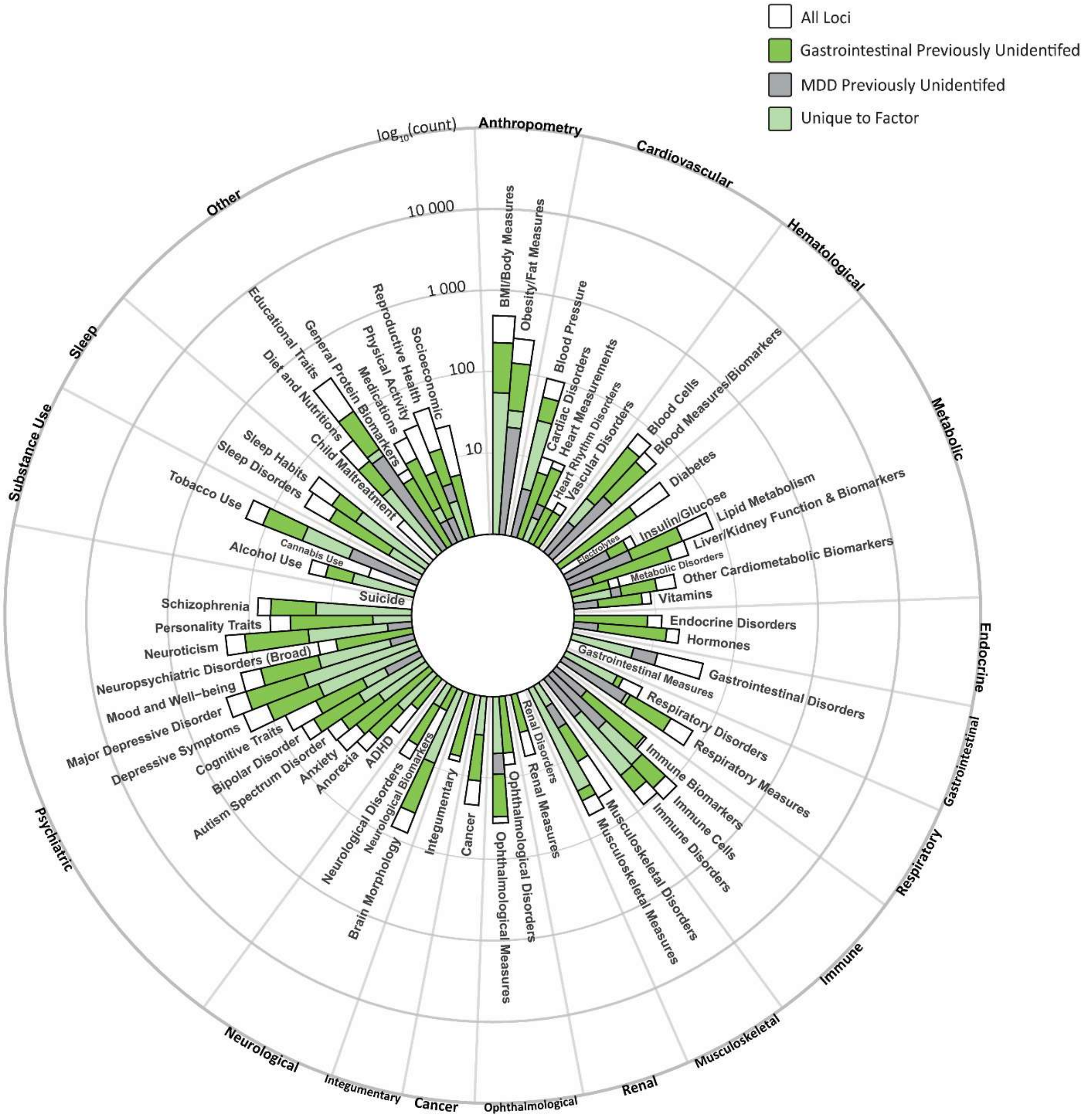

**Supplementary Figure 7c. Phenome-wide association studies of Gastrointestinal-MDD latent factor, linking known disease/trait associations to identified independent loci.** Circular bar plots illustrate the number of associations per trait group on a logarithmic scale. The count of associations is further partitioned into loci previously unidentified to disease traits used in the multivariate GWAS, loci previously unidentified to major depressive disorder (MDD), and loci that are unique to each factor (i.e., loci are not associated with the other disease factors).

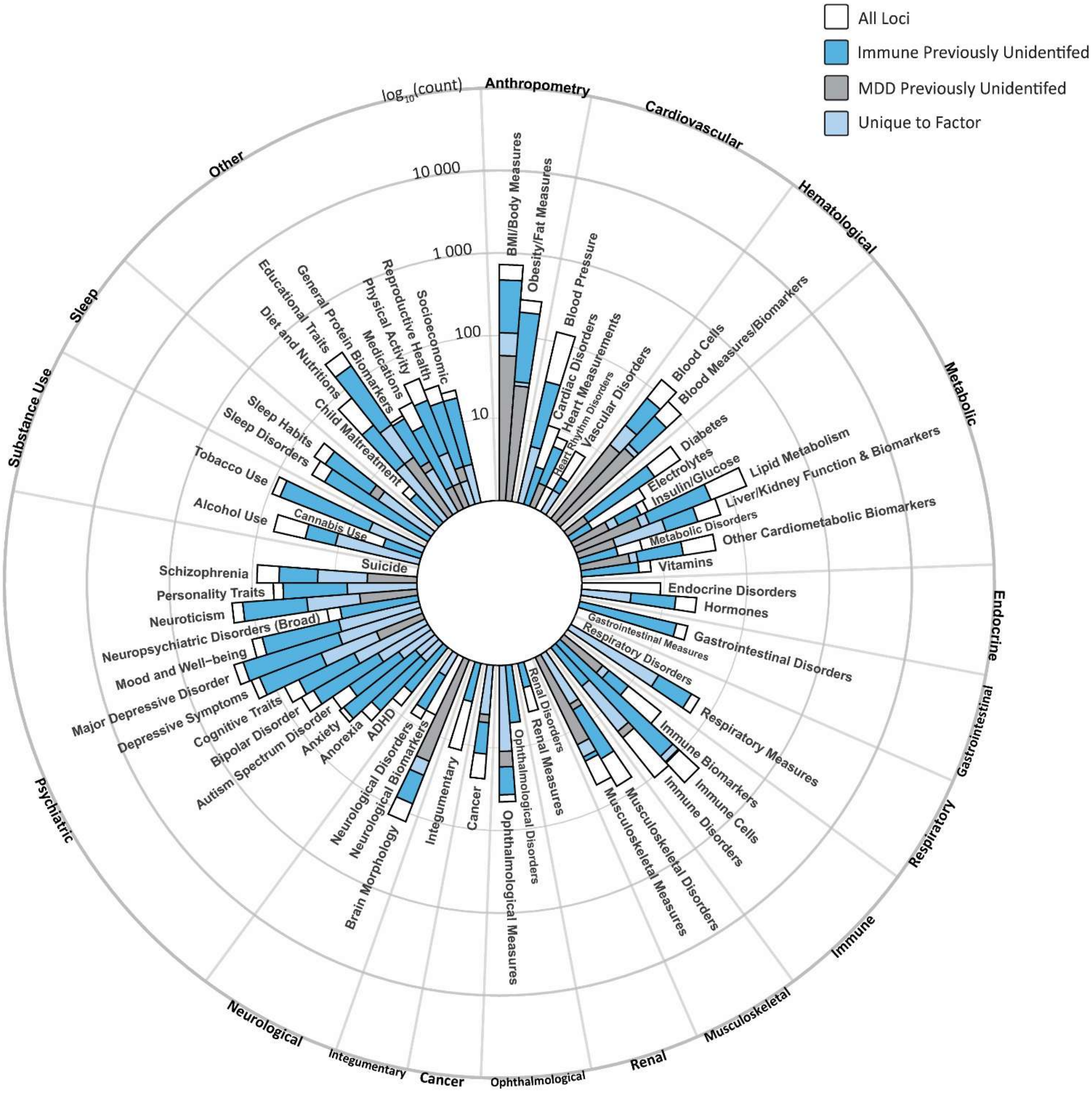

**Supplementary Figure 7d. Phenome-wide association studies of Immune-MDD latent factor, linking known disease/trait associations to identified independent loci.** Circular bar plots illustrate the number of associations per trait group on a logarithmic scale. The count of associations is further partitioned into loci previously unidentified to disease traits used in the multivariate GWAS, loci previously unidentified to major depressive disorder (MDD), and loci that are unique to each factor (i.e., loci are not associated with the other disease factors).

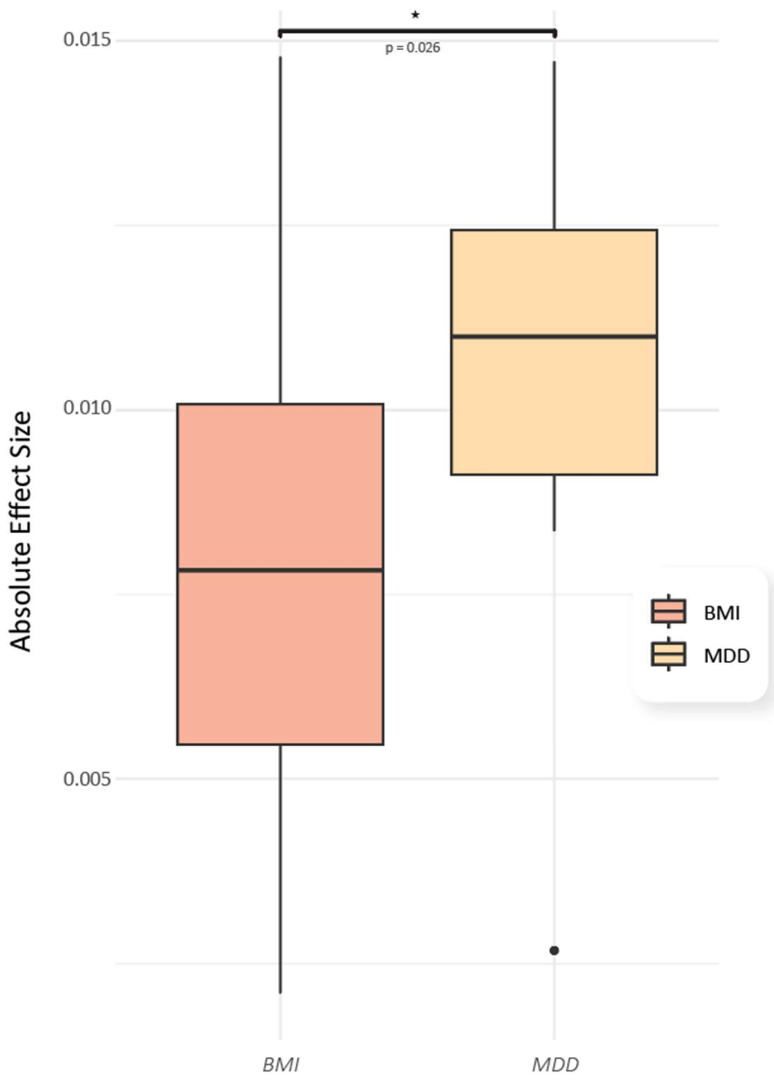

**Supplementary Figure 8. Boxplot comparing absolute effect sizes of gastrointestinal-MDD second-order latent factor variants, selected based on gastro-oesophageal reflux disease (GORD) subgroups.** Genetic variants are associated with depression-driven or obesity-driven GORD loci obtained from Ong et al. (2022). A Wilcoxon test was used to compare the two groups with a star indicating a p-value less than 0.05.

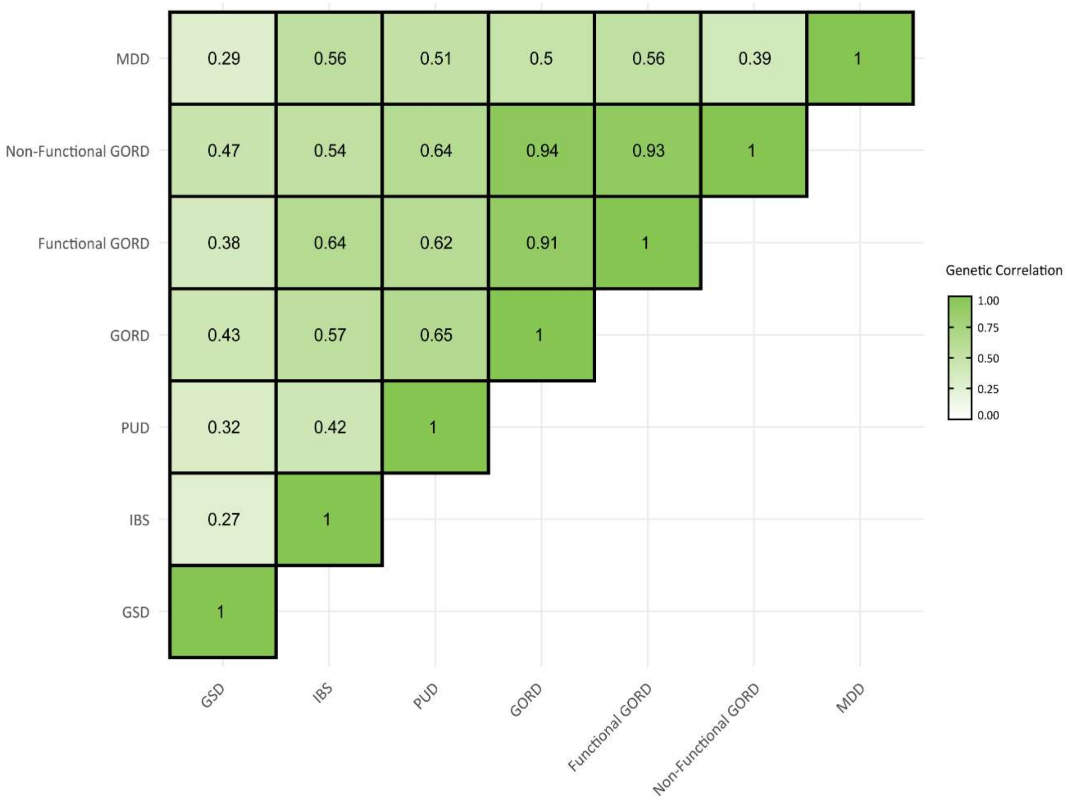

**Supplementary Figure 9. Genetic correlations (rg) between functional and non-functional Gastro-oesophageal Reflux Disease (GORD), Major Depressive Disorder (MDD), and other gastrointestinal traits measured by Linkage Disequilibrium Score Regression estimated from common variants.** Functional and non-functional GORD subtypes were obtained from Ong et al. (2022). Acronyms: Gallstone Disease (GSD), Irritable Bowel Syndrome (IBS), Peptic Ulcer Disease (PUD).

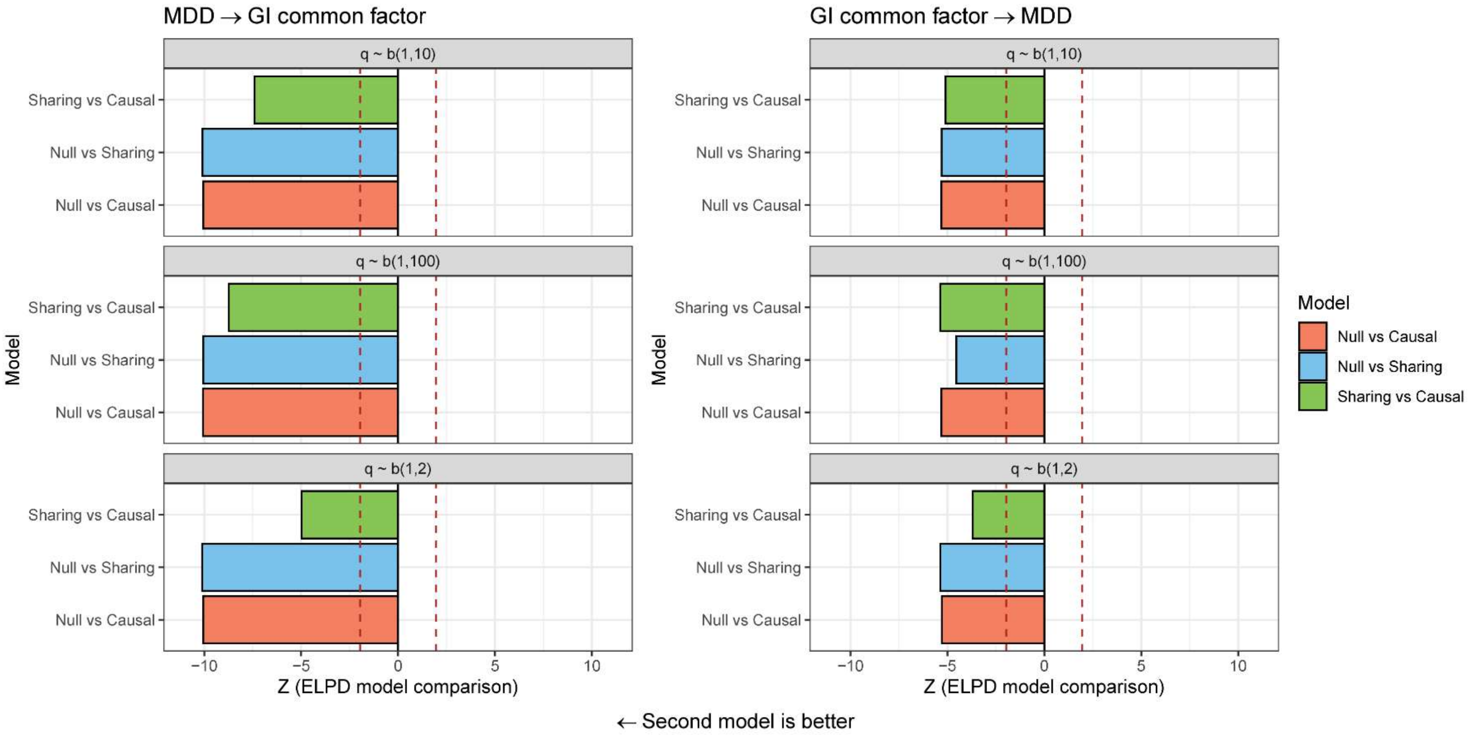

**Supplementary Figure 10. Exploring the relationship between Major Depressive Disorder (MDD) and the gastrointestinal (GI) common factor using the CAUSE model.** We constructed CAUSE models in both directions, MDD to GI common factor (left-hand side), and GI common factor to MDD (right-hand side). The Z score of the change in expected logwise point density (ELPD/standard error of ELPD) is plotted, between the two models in three configurations: null versus sharing, null versus causal, and sharing versus causal. Three different beta priors were implemented: q ~ Beta(1,10), q ~ Beta(1,2), and, q ~ Beta(1,100). The red-dotted lines denote nominal statistical significance ($P < 0.05$). Negative ELPD values imply the second listed model fits better.
